# A machine-learning framework to characterize functional disease architectures and prioritize disease variants

**DOI:** 10.1101/2025.10.23.25338598

**Authors:** Siliangyu Cheng, Artem Kim, Dhrithi Deshpande, Steven Gazal

**Affiliations:** Department of Population and Public Health Sciences, Keck School of Medicine, University of Southern California, Los Angeles, CA, USA; Center for Genetic Epidemiology, Keck School of Medicine, University of Southern California, Los Angeles, CA, USA; Department of Clinical Pharmacy, Alfred E. Mann School of Pharmacy and Pharmaceutical Sciences, University of Southern California, Los Angeles, CA, USA; Department of Quantitative and Computational Biology, University of Southern California, Los Angeles, CA, USA

## Abstract

Modeling disease effect sizes from genome-wide association studies (GWAS) is critical for both advancing our understanding of the functional architecture of human disease and providing informative priors that enhance the prioritization of potentially causal variants. Here, we introduce the variant-to-disease (V2D) framework, an approach that leverages machine-learning algorithms to model disease effect sizes from posterior estimates of effects obtained via genome-wide fine-mapping and functional annotations. We benchmarked the V2D framework using simulations and real data analysis, demonstrating that it provides reliable estimates of heritability (*h*^2^) functional enrichment. By applying the V2D framework with linear trees to 15 UK Biobank traits, we identified non-linear relationships between constraint and regulatory annotations, highlighting constrained regulatory variants as the main functional component of disease functional architecture (*h*^2^ enrichment = 17.3 ± 1.0x across 79 independent GWAS). By applying the V2D framework with neural networks, we developed GWAS prioritization scores, which were extremely enriched in common variant *h*^2^ (20.6 ± 0.7x for the top 1% scores), outperformed existing prioritization scores in the analysis of different GWAS datasets, were transportable to analyze gene expression and non-European datasets, and improved variant prioritization in GWAS fine-mapping studies.

## Introduction

Genome-wide association studies (GWAS) have emphasized that disease-associated variants are extremely enriched in annotations related to distal gene regulation and natural selection ^1–3^. Accurately modeling disease effects from GWAS data as a function of hundreds of these annotations has become central in post-GWAS studies, both for characterizing functional disease architectures ^4–8^ and for providing informative priors that improve the power and accuracy of fine-mapping studies ^9–11^.

The key challenge in this modeling is that GWAS do not directly estimate true disease effects but instead estimate marginal effects that are influenced by linkage disequilibrium (LD) with nearby SNPs. State-of-the-art approaches, such as stratified LD score regression (S-LDSC) ^2,3,7^, have enabled the modeling of disease effect sizes as a linear combination of annotations directly from GWAS marginal statistics. However, linear models are not optimal when continuous annotations do not have a linear relationship with effect sizes or when there are interactions across annotations. Recent approaches have proposed to train machine learning models on variants fine-mapped with high confidence ^12,13^, but it is unclear if these sets are representative of disease polygenic architecture and which non-linear combinations of annotations shape functional disease architectures.

Here, we propose and evaluate the variant-to-disease (V2D) framework, a novel approach that leverages machine-learning algorithms to model disease effect sizes from posterior estimates of squared normalized effect sizes obtained via genome-wide fine-mapping and ∼100 functional annotations (**Fig. 1**). We benchmarked the V2D framework using simulations and real data analysis, demonstrating that it provides reliable estimates of heritability (*h*^2^) functional enrichment. By applying the V2D framework with linear trees, we identified non-linear relationships between annotations related to distal regulation and evolutionary constraint. Finally, we developed V2D GWAS prioritization scores that outperformed existing prioritization scores in *h*^2^ and fine-mapping analyses. In conclusion, we developed a polygenic framework leveraging machine learning models, GWAS and ∼100 functional annotations, which improves both our understanding of disease functional architectures and prediction of causal variants in GWAS.

**Figure 1.**
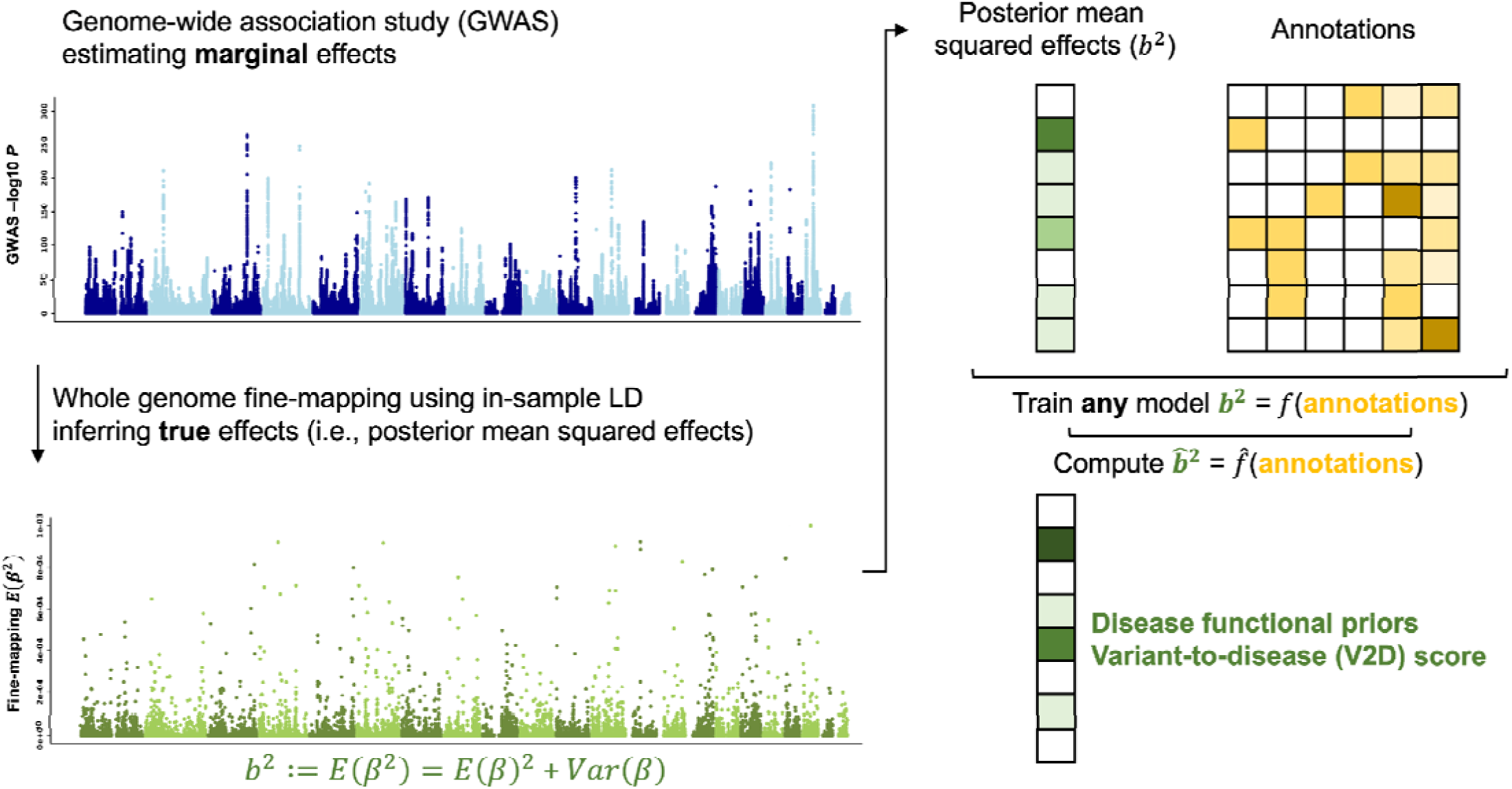
Overview of the variant-to-disease (V2D) modeling framework. The V2D framework leverages genome-wide estimates of posterior mean squared causal effect sizes ^2^ and functional annotations to predict ^2^, defined as V2D scores. We estimated ^2^ for 15 independent UK Biobank traits using PolyFun and SuSiE and averaged normalized values across traits.

## Results

### Overview of the V2D framework

We define *β*_j_ as the normalized effect size of SNP *j* on a given trait. Because of LD between variants, these effects cannot be directly measured in GWAS, which prevents the direct modeling of disease effects on functional annotations. The intuition of the V2D framework is that it leverages the posterior mean estimates of *β* obtained from fine-mapping methods to perform such modeling (**Fig. 1**). This framework consists of three steps. First, genome-wide fine-mapping is performed to estimate the posterior mean E(*β*_j_) and variance Var(*β*_j_) for all variants. We define posterior mean squared causal effect sizes as *b*_j_^2^ := E(*β*_j_)^2^ *+* Var(*β*_j_). Second, for a given machine learning method, optimal hyperparameters are identified by minimizing the mean squared error (MSE) under a leave-odd/even-chromosome-out (LEOCO) cross-validation scheme. Third, the model uses optimal hyperparameters to predict *b̂*^2^ under the LEOCO scheme; these predictions are defined as V2D scores.

Here, we modeled *b*^2^ estimated across 15 independent UK Biobank GWAS (average *N* = 297K; **Table S1**) for ∼20M SNPs with MAF ≥ 0.1%. For each trait, *b*^2^ estimates were obtained by PolyFun ^11^ using the default SuSiE with uniform priors (i.e., no functional priors; SuSiE-noprior) and with functional priors obtained from a linear combination of the annotations of the baseline-LF model as estimated by S-LDSC (SuSiE-prior); results obtained with SuSiE-noprior and SuSiE-prior are compared throughout the manuscript. We assigned to each SNP *j* the mean value of normalized of *b*_j_^2^ (i.e., *b*_j_^2^/*Σ b* ^2^) across the 15 traits and modeled these values using the 187 annotations of the baseline-LF model ^3,7^ (**Table S2**). Analyses were run independently on common (MAF≥ 5%) and low-frequency (0.1% ≤ MAF < 5%) variants to account for the effect of negative selection within annotations ^14^ (e.g., by giving a higher weight to non-synonymous variants in low-frequency functional architecture). We investigated four popular machine learning models (decision tree, multi-layer perceptron (MLP) neural networks ^15^, extreme gradient boosting (XGBoost) ^16^, and random forests ^17^), as well as a linear model as a baseline. We evaluated V2D scores across datasets selected to minimize their dependence on the 15 UK Biobank GWAS; we notably computed *h*^2^ enrichment across 79 independent European GWAS not correlated with the 15 UK Biobank traits (**Table S3**).

Further details are provided in **Methods**. We have released open-source software implementing our framework (see **Code availability**) and have made all V2D scores for ∼20M UK Biobank SNPs with MAF >0.1% publicly available (see **Data availability**).

### Evaluating posterior mean squared causal effect sizes estimates from SuSiE using simulations

We performed simulations to evaluate how *b*^2^ estimates from SuSiE provide accurate characterization of disease functional architectures. For each simulation, we ran SuSiE with an in-sample LD and no functional priors (SuSiE-noprior) as well as priors estimated on an independent simulation (SuSiE-prior). For each simulation scenario, we considered three levels of trait heritability *h^2^* (0.5, 0.2, and 0.1). We evaluated SuSiE *b*^2^ estimates by comparing the true *h*^2^ functional enrichment of annotations obtained from simulated *β*^2^ (i.e., the mean *β*_j_^2^ for all variants within an annotation divided by the mean *β*_j_^2^ for all variants) to those obtained from estimated *b*^2^. Further details are provided in **Methods**.

First, we simulated causal effect sizes as a linear combination of functional annotation (**Fig. 2A** and **Table S4**). We considered the annotations from the baseline-LD model and obtained their corresponding linear coefficients by averaging their S-LDSC estimates obtained across the 15 independent UK Biobank traits. We then evaluated SuSiE functional enrichment estimates for 40 main annotations of the baseline-LD model (**Table S2**). SuSiE-prior leveraged priors estimated from a linear model regressing effects from an independent simulation on all annotations of the baseline-LD model. For simulations under *h^2^* = 0.5, both SuSiE-noprior and SuSiE-prior produced nearly unbiased enrichments (*r* > 0.99; slopes = 0.96 and 1.01, respectively). For lower values of *h^2^*, both approaches maintained a high correlation between simulated and estimated enrichments across annotations (*r* > 0.99), but SuSiE-noprior tended to underestimate enrichments (slope = 0.82 and 0.67 for *h^2^* = 0.1 and 0.05, respectively), whereas SuSiE-prior tended to slightly overestimate enrichments (slope = 1.06 and 1.09, respectively).

**Figure 2.**
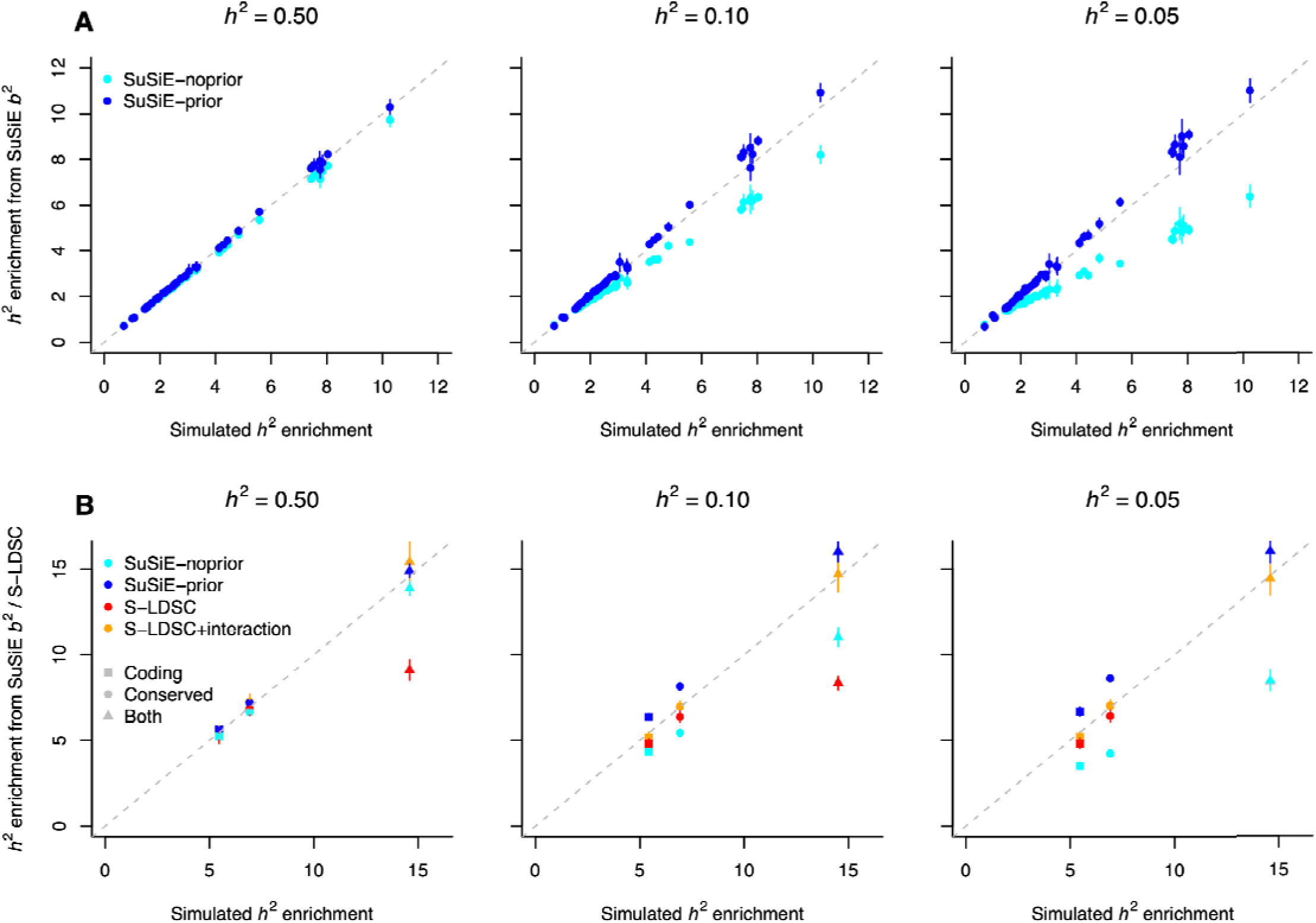
Evaluating SuSiE *b*^2^ estimates using simulations. We report heritability (*h*^2^) enrichment estimates from SuSiE with no priors (SuSiE-noprior) and with priors (SuSiE-prior) estimated from independent simulations (SuSiE-prior). (**A**) Simulations were performed using per-SNP *h^2^* from the baseline-LD model, and SuSiE enrichments were reported for 40 representative functional annotations. (**B**) Simulations incorporating an interaction between coding and conserved annotations were analyzed with SuSiE and S-LDSC, both without and with the interaction term (S-LDSC and S-LDSC+interaction, respectively). Error bars represent 95% confidence intervals. Numerical results are reported in **Table S4** and **Table S5**.

Second, we simulated causal effect sizes using a non-linear combination of functional annotations, to investigate scenarios in which the linear model used by SuSiE-prior does not match the generative model (**Fig. 2B** and **Table S5**). We considered a model in which E[*β*_j_^2^] depends only on the coding and conserved annotations, with the expected effect being higher when a variant is both coding and conserved (coding&conserved). We evaluated SuSiE enrichments for coding, conserved and coding&conserved variants. SuSiE-prior leveraged priors estimated from a linear model by regressing effects from an independent simulation on the coding and conserved annotations only. We also evaluated S-LDSC estimates considering only the coding and conserved annotations (as is typically done in practice) as well as estimates including an additional explicit interactive term (S-LDSC+interaction). For *h^2^*= 0.5, all models produced nearly unbiased estimates of enrichment for the coding and conserved annotations. For coding&conserved variants, SuSiE and S-LDSC+interaction estimates were nearly unbiased, whereas S-LDSC showed a strong downward bias. This result suggests that SuSiE *b*^2^ estimates can capture characterization of functional architectures that would be missed by a purely linear model. For lower values of *h^2^*, SuSiE-noprior underestimated enrichments for coding&conserved variants, whereas SuSiE-prior overestimated them but with smaller absolute bias. S-LDSC+interaction yielded unbiased estimates for all annotations and across all values of *h^2^*, highlighting that S-LDSC can be leveraged to validate *h^2^* enrichment of non-linear combinations of annotations.

Altogether, these simulations demonstrate that SuSiE *b* ^2^ estimates can accurately characterize functional architectures in well-powered GWAS and identify non-linear combinations of annotations. For less-powered GWAS, using priors from a linear model improved (but slightly overestimated) *b* ^2^ estimates. Therefore, we decided to leverage *b* ^2^ using functional priors from the (additive) baseline-LF model in downstream V2D analyses (results from *b*^2^ using no priors are presented in Supplemental material) and to validate *b*^2^ estimates and V2D predictions via *h^2^* analyses with S-LDSC.

### Benchmarking SuSiE *b^2^* estimates on 15 UK Biobank traits

We benchmarked our *b*^2^ estimates averaged across 15 UK Biobank traits by comparing their *h^2^* enrichment on representative annotations to S-LDSC estimates averaged across the same traits (used here as a gold standard). Consistent with our simulations, SuSiE-noprior enrichments for the 40 main annotations were highly correlated but were consistently lower than those from S-LDSC (*r* = 0.83, slope = 0.53). In contrast, SuSiE- prior enrichments were highly correlated but slightly higher than those from S-LDSC (*r* = 0.96, slope = 1.09) (**Fig. 3** and **Table S6**). SuSiE enrichments for the non-synonymous annotation were significantly higher than those from S-LDSC, likely because non-synonymous variants have larger effect sizes and are downweighted as outliers by S-LDSC (**Fig. S1**). Also, SuSiE enrichments for SNPs in different LD score bins were similar to those obtained with S-LDSC (**Fig. S2** and **Table S7**). Of note, variants fine-mapped with high confidence (posterior inclusion probability (PIP) > 0.90, as performed in refs. ^12,13^) tended to be disproportionately enriched for variants within the lowest LD bin (2.82x vs. 1.32x with S-LDSC), likely because such variants are easier to fine-map; similar patterns were observed for variants fine-mapped in other GWAS and expression quantitative trait loci (eQTL) datasets (**Fig. S3**). Hence, leveraging *b*^2^ estimates from fine-mapping analyses (rather than PIP) may provide a more balanced model of effect sizes that does not overweight low-LD variants. Finally, we replicated our conclusions using S-LDSC results meta-analyzed across 79 independent GWAS (**Fig. S4**), demonstrating that our *b*^2^ estimates across 15 UK Biobank traits are representative of the broader genetic architecture of human diseases and complex traits.

**Figure 3.**
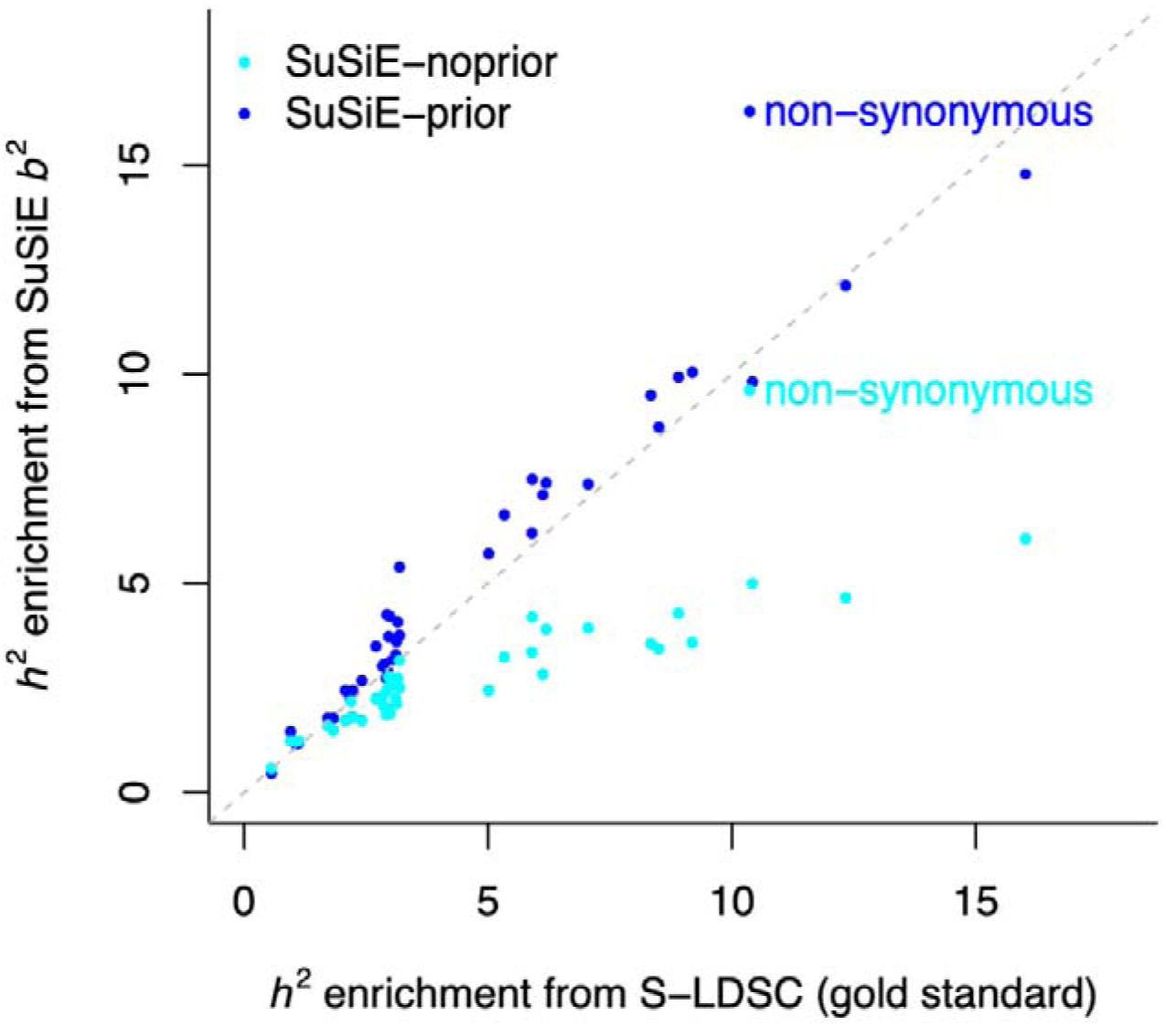
Evaluating SuSiE *b*^2^ estimates on 15 UK Biobank traits. We report estimates of *h^2^* enrichment estimated across 15 UK Biobank traits with S-LDSC (used as the gold standard), SuSiE-noprior, and SuSiE-prior (priors were obtained with PolyFun) for 40 main functional annotations from the baseline-LD model. Numerical results and corresponding standard errors are reported in **Table S6**.

### Leveraging decision trees to visualize non-linear relationships of functional annotations

To demonstrate the presence of non-linear relationships between functional annotations and variant effects, we applied decision trees (because they provide interpretable visualization of complex relationships) to SuSiE- prior *b*^2^ estimates and annotations of the baseline-LF model (**Fig. 4**). Common and low-frequency SNPs were analyzed separately. We defined the optimal hyperparameters of the decision trees using a LEOCO scheme and observed that MSE was minimized when using a leaf size of 25K common SNPs and 10K low-frequency SNPs and did not greatly improve nor overfit after a depth > 7 (**Figs. 4A-4B** and **Table S8**). Notably, more than one-third of nodes (10/27 for common SNPs and 16/36 for low-frequency SNPs) corresponded to continuous annotations (e.g., CpG content, allele age) for which the regression tree was able to identify an optimal cut-off (**Figs. S5-S6**). Similar conclusions were obtained with SuSiE-noprior *b* ^2^ estimates (**Figs. S7-S8**). For interpretability, we visualized trees of depth 3 **(Figs. 4C–D)** and describe their main features below.

**Figure 4.**
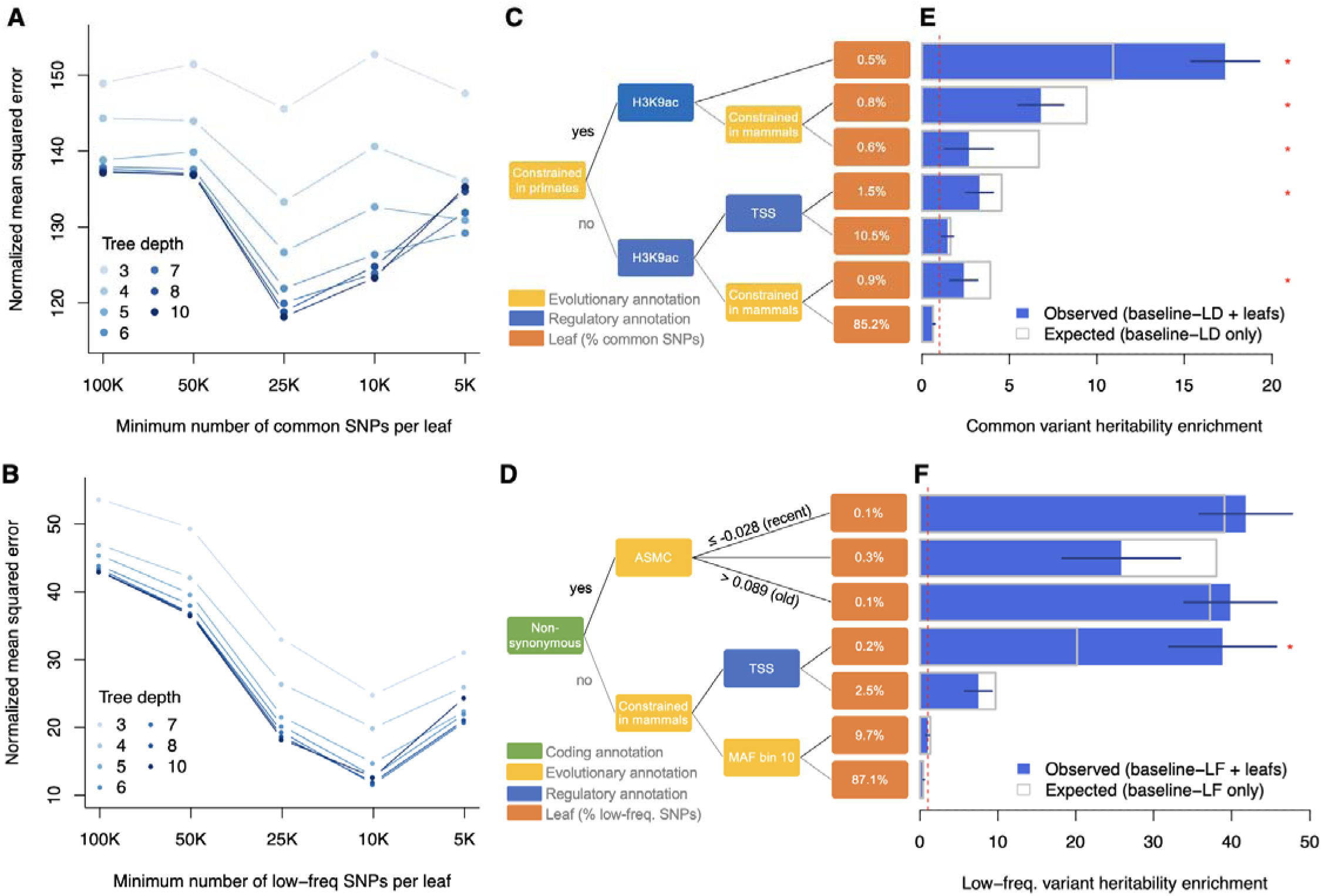
Visualizing non-linear relationships of functional annotations with decision trees. **(A,B)** We report normalized mean squared errors (MSEs) of decision trees with varying depth and minimum SNPs per leaf, estimated using a LEOCO approach for common (A) and low-frequency (B) SNPs. Errors are normalized by the MSE from the linear model. Numerical results are reported in **Table S8**. **(C,D)** We report decision trees of depth 3 based on SuSiE-prior _j_^2^ estimates averaged across 15 UK Biobank traits for common (**C**) and low-frequency (**D**) SNPs. Trees of depth 7 are shown in **Figs. S5-S6**. **(E,F)** We report the common variant *h^2^* enrichment of each leaf estimated by S-LDSC (blue) and expected by S-LDSC with the baseline-LD model (grey) using 79 independent European GWAS not correlated with the 15 UK Biobank traits (**E**). We report the low-frequency variant *h^2^*enrichment of each leaf estimated by S-LDSC (blue) and expected by S-LDSC with the baseline-LF model (grey) using 27 independent UK Biobank traits with sufficient power to investigate low-frequency variant architecture (**F**). Error bars represent 95% confidence intervals. Asterisks represent significant differences (*P* < 0.05/7) between observed and expected enrichments. Numerical results are reported in **Tables S9** and **S10**. TSS: transcription start site; ASMC: ascertained sequentially Markovian coalescent (values normalized and adjusted to minor allele frequency) ^18^.

For common SNPs, the leaf with the highest mean SuSiE-prior *b*_j_^2^ consists of SNPs that are both constrained and in H3K9ac peaks, which points to the role of constrained regulatory variants into common variant functional architectures (**Fig. 4C** and **Fig. S4**). To quantify the informativeness of our trees across an independent set of traits, we created one SNP annotation per leaf of the common SNP tree and ran S-LDSC with the baseline-LD model across 79 independent traits (**Fig. 4E** and **Table S9**). SNPs in the main leaf (i.e., constrained and in H3K9ac peaks) were highly enriched in *h^2^* (0.5% of common SNPs explaining 9.0 ± 0.5% of *h^2^*, enrichment = 17.3 ± 1.0x). Overall, 6 of 7 leaves were significantly (*P* < 0.05/7) enriched or depleted of *h^2^*, which supports the generalizability of this approach. To evaluate whether these leaves highlight interactive effects across annotations, we compared the *h^2^*estimated by S-LDSC when integrating leaf annotations to the one expected from the baseline-LD model (i.e., without leaves; **Fig. 4E**). The main leaf showed significantly higher enrichment than expected (17.3 ± 1.0x vs. 10.9 ± 0.5x; *P* = 1.4 x 10^-8^), indicating the baseline-LD model underestimates the effect sizes of constrained regulatory variants. For 4 of the remaining 6 leaves, a model without the leaves overestimated their *h^2^* enrichment. Similar results were obtained with deeper trees (8 of 17 leaves at depth 5 showed significant differences) and with SuSiE-noprior *b*^2^ estimates (**Fig. S9**).

For low-frequency SNPs, the first split corresponded to non-synonymous SNPs, consistent with distinct architectures between common and low-frequency SNPs ^14^ (**Fig. 4D** and **Fig. S6**). Non-synonymous SNPs were further split by predicted allele age ^18^, with higher mean SuSiE-prior *b*_j_^2^ for variants with the most recent and most ancient age, thus suggesting a balance between different evolutionary forces on non-synonymous variants. The tree also highlighted high mean SuSiE-prior *b*_j_^2^ for constrained variants in transcription starting sites (TSS). To validate these results, we created one SNP-annotation per leaf and applied our extension of S-LDSC to low-frequency variants with the baseline-LF model across 27 independent UK Biobank traits with sufficient power to analyze low-frequency variant architectures ^14^ (**Fig. 4F** and **Table S10**). We confirmed that both the most recent and most ancient non-synonymous low-frequency variants were more enriched in *h^2^* than other non-synonymous low-frequency variants (one-sided *P* = 5.7 x 10^-4^ and 2.4 x 10^-3^, respectively), a pattern not predicted by the baseline-LF model. Constrained variants at TSS were also significantly more enriched for low-frequency variant *h^2^* than expected under the baseline-LF model (38.9 ± 3.5x vs. 20.2 ± 1.4x; *P* = 7.1 x 10^-7^), highlighting that constrained regulatory variants are also a significant component of low-frequency variant functional architectures.

Altogether, these results illustrate the advantages of the V2D framework: 1) it reveals the key role of constrained regulatory variants in shaping the common and low-frequency variant functional architectures of human diseases, 2) it successfully reveals non-linear and interactive effects between functional and evolutionary annotations that traditional linear models fail to capture, and 3) it demonstrates generalizability to traits not genetically correlated with those used for training.

### Creating V2D scores enriched in disease heritability and fine-mapped variants

We applied the V2D framework to common variants with more complex machine learning methods to create V2D scores and evaluated their disease informativeness using three GWAS datasets chosen to minimize dependence on the 15 UK Biobank GWAS used for training: the 79 independent European GWAS, common variants fine-mapped with high confidence (PIP > 0.90) in 931 GWAS from the Million Veteran Program (MVP) ^13,19^, and common variants fine-mapped with high confidence in 1,248 GWAS from FinnGen ^20^. For each dataset, we computed *h^2^*enrichment or excess overlap at five score thresholds, from the top 0.2% to the top 5% of variants. V2D scores obtained with the MLP neural network outperformed those from the linear model, random forest and XGBoost in at least two of the three datasets (**Fig. 5A** and **Table S11**; hyperparameters used for each model are reported in **Table S12**). Training scores with SuSiE-noprior *b*^2^ estimates yielded lower *h^2^* enrichment (though similar excess overlap), which confirms that functional priors improve disease effect prediction (**Fig. S10**). For downstream analyses, we leveraged the V2D scores obtained by the MLP neural network and labeled these scores V2D-MLP. All V2D-MLP scores for ∼20M of UK Biobank SNPs with MAF > 0.1% are publicly available (see **Data availability**).

**Figure 5.**
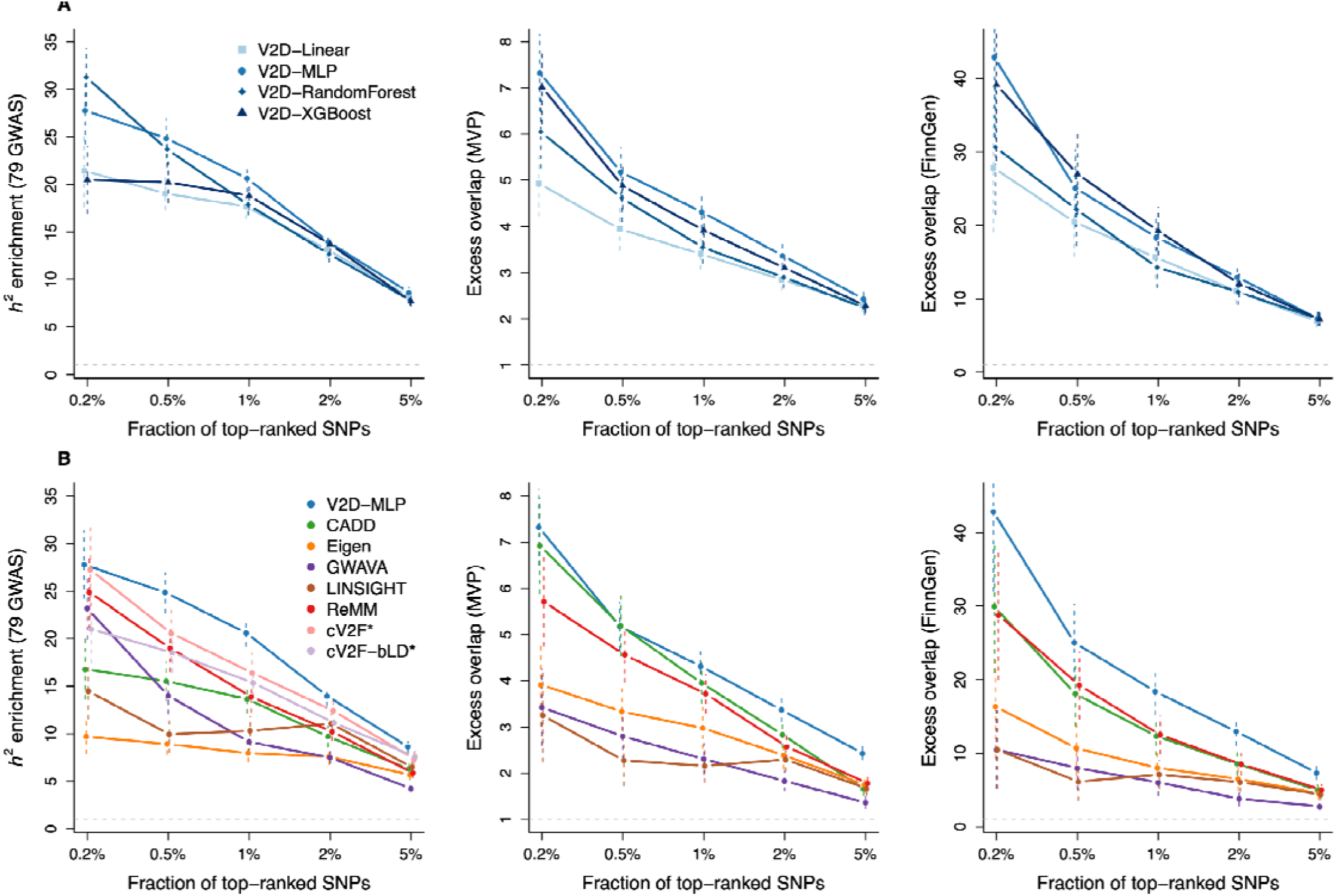
Benchmarking V2D scores across machine learning models and existing prioritization scores. **(A)** We report *h^2^* enrichment computed across 79 independent GWAS and excess overlap computed across common variants fine-mapped with high confidence in MVP and FinnGen for V2D scores obtained with different machine learning methods. **(B)** Comparable benchmarks for V2D-MLP against existing prioritization methods. Error bars represent 95% confidence intervals. Numerical results are reported in **Tables S11** and **S13**. * cV2F and cV2F-bLD were not evaluated with fine-mapped datasets, because cV2F scores are specifically trained on variants fine-mapped with high confidence and are inherently enriched toward SNPs with low LD versus other prioritization scores (**Fig. S3** and **Fig. S11**).

We next compared V2D-MLP scores to six state-of-the-art prioritization scores (CADD ^21^, Eigen ^22^, GWAVA ^23^, LINSIGHT ^24^, ReMM ^25^ and cV2F ^13^) as well as cV2F scores re-computed on the same baseline-LD annotations as V2D-MLP (cV2F-bLD). V2D-MLP scores provided higher *h^2^* enrichment and higher or similar excess overlap than all other scores (**Fig. 5B** and **Table S13**). Specifically, the top 1% of V2D-MLP scores had an *h^2^*enrichment of 20.6 ± 0.7x, which was significantly higher than those obtained by cV2F (16.4 ± 1.1x; *P* = 0.001 for difference) and ReMM (13.9 ± 0.8x; *P* = 8.7 x 10^-10^ for difference). The top 2% and 5% of V2D-MLP scores also showed significantly higher excess overlap with MVP and FinnGen fine-mapped variants than those from other methods. Because cV2F scores are ascertained to identify variants fine-mapped with high confidence (because they are used for training), they are inherently enriched in SNPs with low LD as compared with other prioritization scores (**Fig. S11**); for this reason, cV2F and cV2F-bLD were not evaluated on the fine-mapped datasets (also enriched in SNPs with low LD, **Fig. S3**). We observed high excess overlap on fine-mapped eQTL from eQTLGen ^26,27^ as well as 233K variants functionally tested using massively parallel reporter assay (MPRA) across 5 cell lines ^28^ (**Fig. S12A**), which demonstrates the transportability of V2D-MLP scores across gene expression phenotypes. We replicated these findings using *h^2^*enrichment from 20 independent East-Asian GWAS ^29–31^ and excess overlap with fine-mapped variants from 79 Biobank Japan GWAS ^32^ and fine-mapped eQTLs from the Japan COVID-19 Task Force (JCTF) ^33^ (**Fig. S12B**), which demonstrates the transportability of V2D-MLP scores across non-European populations.

Overall, V2D-MLP scores outperformed those from existing prioritization methods in GWAS analyses and demonstrated strong transportability across both gene expression and non-European datasets.

### Leveraging V2D scores to prioritize disease variants

A key feature of V2D scores is that they are proportional to *β*^2^ and can be directly leveraged as priors for fine-mapping ^11^. Therefore, we used V2D-MLP scores to perform functionally informed fine-mapping of 110 UK Biobank traits by weighting posterior probabilities of SuSiE credible sets (CS), leveraging the frameworks of refs. ^12,13^ (method labeled SuSiE+V2D). Using SuSiE+V2D, we fine-mapped 4,265 (resp. 5,704 and 8,854) variants with PIP > 0.90 (resp. 0.75 and 0.50) when using SuSiE+V2D, which represents an increase of 20.9% (resp. 28.7% and 31.3%) from SuSiE fine-mapping results (**Fig. 6A**, **Tables S14** and **S15**). To test whether SuSiE+V2D better prioritizes causal variants than SuSiE, we examined 3,323 CS in which the top-PIP variant differed between the two methods and variants had been functionally tested by MPRA in ref. ^28^. Variants prioritized by SuSiE+V2D were nearly twice more likely to be expression-modifying variants (emVars) than those prioritized by SuSiE (20.8 ± 0.7% vs 10.7 ± 0.5%, *P* = 1.3 x 10^-29^; **Fig. 6B** and **Table S16**). We next compared SuSiE+V2D with functionally-informed fine-mapping approaches using cV2F and cV2F-bLD scores (SuSiE+cV2F and SuSiE+cV2F-bLD, respectively). SuSiE+V2D identified 4.3% (resp. 3.6% and 4.4%) more variants fine-mapped with PIP > 0.90 (resp. 0.75 and 0.50) than SuSiE+cV2F and 4.9% (resp. 5.5% and 6.8%) more variants than SuSiE+cV2F-bLD (**Fig. 6A** and **Table S15**). Variants prioritized by SuSiE+V2D were more likely to be emVars than those prioritized by SuSiE+cV2F-bLD (18.3 ± 0.8% vs. 13.3 ± 0.7% across 2,519 CS, *P* = 4.9 x 10^-7^; **Fig. 6C** and **Table S17**); SuSiE+cV2F was not evaluated so as to avoid circularity because it incorporates the same MPRA dataset into its training features.

**Figure 6.**
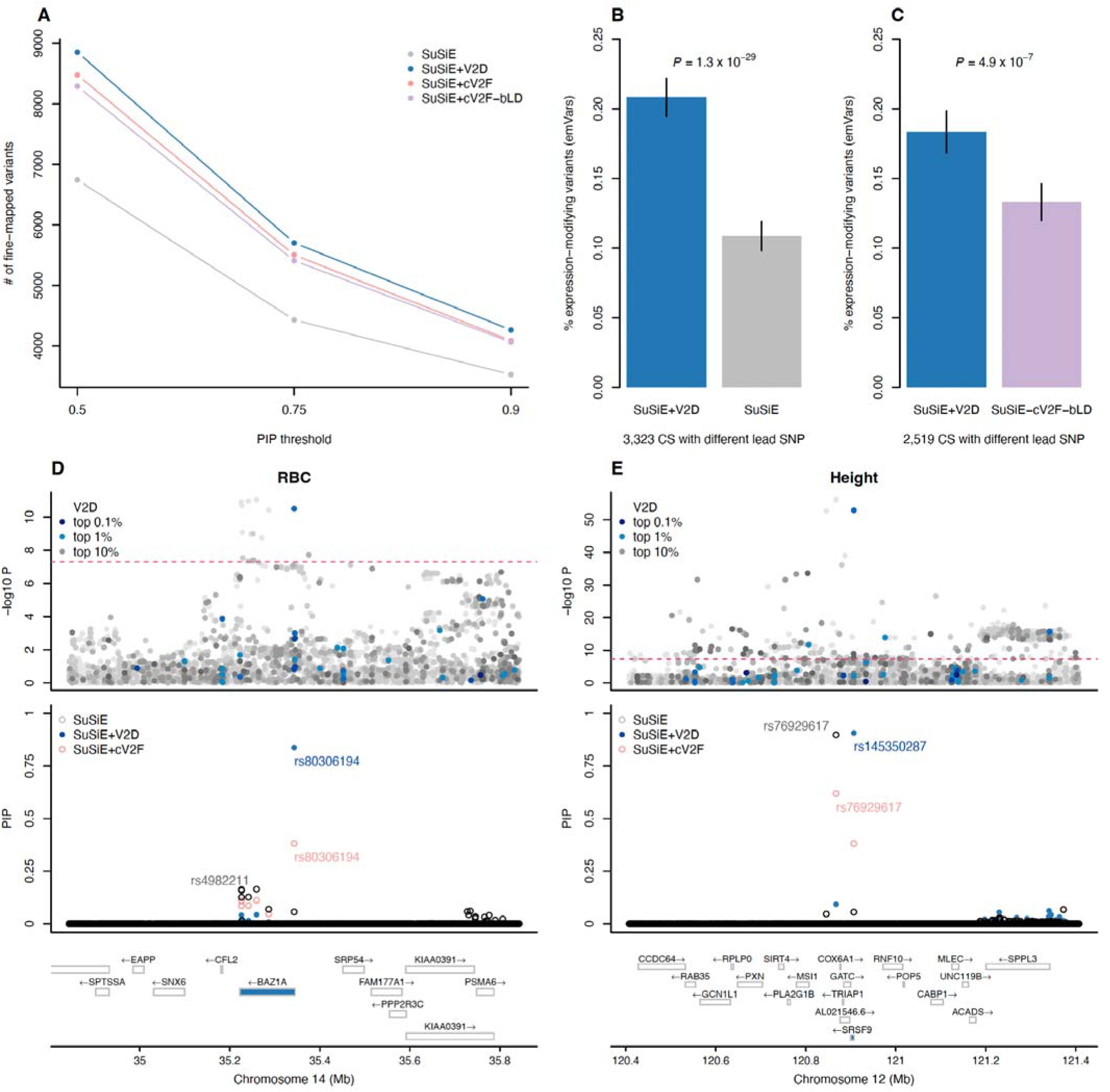
Leveraging V2D scores to prioritize disease variants. **(A)** We report the number of fine-mapped SNPs exceeding posterior inclusion probability (PIP) thresholds across 110 UK Biobank traits. Numerical results are reported in **Table S15**. **(B,C)** We report the fraction of lead SNPs (variants with the highest PIP) that are expression-modifying variants (emVars) in credible sets with different lead SNPs from SuSiE and SuSiE+V2D (**B**) and from SuSiE+V2D and SuSiE+cV2F-bLD (**C**). Massively parallel reporter assay (MPRA) results are reported in **Tables S16** and **S17**. (**D,E**) We report two examples where SuSiE+V2D was able to prioritize a variant with at least twice more confidence than other fine-mapping approaches for red blood cell count (RBC) (**D**) and height (**E**). We plotted the −log10 GWAS *P* values of each SNP (top), PIP values (middle; highlighted are the SNPs with the highest PIP of each method) and the genes in the locus (bottom; closest gene of the variant highlighted by SuSiE+V2D is in blue).

We highlight two examples in which SuSiE+V2D prioritized a variant with at least twice the confidence of other approaches, enabling nomination of more likely functional variants and target genes. At the *BAZ1A* locus in the red blood cell count GWAS, SuSiE+V2D prioritized the constrained regulatory variant rs80306194 (as defined in **Fig. 4C**) with PIP = 0.84 as compared with 0.06 and 0.38 for SuSiE and SuSiE+cV2F, respectively (**Fig. 6D**). All eight SNPs in this CS were tested by MPRA ^28^, and only rs80306194 was identified as an emVar in K562 (**Fig. S13**), which provides additional evidence that rs80306194 is the causal variant at this locus. In the height GWAS, SuSiE+V2D prioritized the *SRSF9* missense variant rs145350287 with high confidence (PIP = 0.91; **Fig. 6E**), which suggests the role of *SRSF9* (a splicing factor) in height etiology. In contrast, SuSiE and SuSiE+cV2F would have prioritized rs76929617 (PIP = 0.90 and 0.62, respectively), making it more challenging to nominate *SRSF9* (6^th^ closest gene) as a candidate gene. Notably, rs145350287 replicated in MVP and FinnGen height fine-mapping studies with PIP = 0.99 and 0.94, respectively.

Finally, we investigated whether functionally informed fine-mapping could be improved by integrating trait-relevant annotations in our V2D-MLP scores. To achieve this, we created trait-specific V2D-MLP scores by using a simple linear combination of V2D scores, trait-relevant cell-type-specific annotations identified by CT-FM ^34^, and their interaction; linear coefficients were estimated by using S-LDSC with a LEOCO scheme from the trait summary statistics (see **Methods**). To validate our approach, we computed these trait-specific V2D scores for 45 of the 79 independent European GWAS for which CT-FM had identified causal cell-type-specific annotations (1.24 annotations per trait on average). The top scores of these trait-specific V2D scores were more enriched in *h*^2^ than were V2D-MLP scores (19.1 ± 0.9x vs. 14.6 ± 0.6x for the top 2% of the scores; *P* = 5.7 x 10^-5^) (**Fig. S14**). Next, we created trait-specific V2D scores for seven well-powered blood-related UK Biobank traits that had SuSiE and CT-FM results available (mean of 1.29 annotations per trait). By leveraging these scores with SuSiE (SuSiE-V2Dx), we could fine-map 45.9% (resp. 53.1% and 52.0%) more variants with PIP > 0.90 (resp. 0.75 and 0.50) than SuSiE, and between 21.9% and 47.3% more variants at these PIP thresholds than SuSiE+V2D, SuSiE+cV2F, and SuSiE leveraging blood-specific cV2F scores (**Fig. S15**). Importantly, variants prioritized by SuSiE-V2Dx were significantly more likely to be emVars than variants prioritized by SuSiE (27.2 ± 1.5% vs. 13.5 ± 1.1% across 903 discordant CS; *P* = 6.1 x 10^-13^) and SuSiE+V2D (26.1 ± 2.4% vs. 16.2 ± 2.0% across 314 discordant CS; *P* = 0.005); similar results were observed with K562 emVars (**Fig. S16**). Lastly, applying SuSiE+V2Dx to red blood cell count and height increased the PIP of rs80306194 and rs145350287 to values > 0.99. Hence, functionally informed fine-mapping can be effectively improved by integrating V2D-MLP scores with both trait-relevant cell type-specific annotations and trait summary statistics.

Overall, V2D-MLP scores enabled fine-mapping variants with high confidence and high functional evidence, allowing to prioritize variants and genes in GWAS follow-up studies.

## Discussion

We developed the V2D framework, a polygenic approach leveraging machine learning models to improve both our understanding of disease functional architectures and prediction of causal variants in GWAS. This framework represents a substantial advance over methods that rely solely on linear combinations of annotations or on variants fine-mapped with high confidence. In addition, V2D scores are proportional to disease square effect sizes and can be directly leveraged as priors for fine-mapping. Using decision trees, we identified non-linear relationships between constraint and regulatory annotations, thus highlighting constrained regulatory variants as a central component of disease architecture (*h*^2^ enrichment = 17.3 ± 1.0x for constrained common variants in H3K9ac peaks and 38.9 ± 3.5x for constrained low-frequency variants in TSS). These analyses also suggested a balance of evolutionary forces acting on low-frequency non-synonymous variants. By applying the V2D framework with MLP neural networks, we constructed GWAS prioritization scores (V2D- MLP) that were greatly enriched in common variant *h*^2^ (enrichment of the top 1% scores = 20.6 ± 0.7x), outperformed existing prioritization methods across GWAS datasets, were generalized to gene expression and non-European data, and improved variant prioritization in fine-mapping studies (notable examples include the *BAZ1A* locus in the red blood cell GWAS and the *SRSF9* locus in the height GWAS). Finally, we introduced a straightforward framework to compute trait-specific V2D-MLP scores.

Our findings have several implications for downstream analyses. First, they highlight the value of posterior mean squared causal effect sizes from fine-mapping, which enable complex modeling of disease effects without explicitly accounting for LD. We recommend using these estimates rather than variants fine-mapped with high confidence in future studies because they tend to be disproportionately enriched for variants with low LD and thus might not be representative of disease LD-dependent architectures (**Fig. S3**). Second, we revealed interactive effects between functional and evolutionary annotations. In particular, constraint regulatory variants emerged as primary drivers of the disease functional architecture, which highlights the need to identify those variants by integrating recent estimates of constraint from different time scales (e.g., mammals ^31,34^, primates ^35^, humans ^36,37^) with functional genomics data. The advantage of the V2D framework is that it can leverage continuous values (e.g., phyloP/phastCons scores for conservation, accessibility score or coverage value from functional assays) to optimally identify such variants. Our results also highlight the role of ancient low-frequency non-synonymous variants. Although more recent variants are more likely to be deleterious because of negative selection ^3^, our results suggest that ancient non-synonymous variants may also have deleterious effects. This pattern could result from different evolutionary pressures, such as balancing selection, or to recent changes in the environment, such as changes in diet or the impact of infectious diseases. More research is needed to fully understand this pattern. Third, although we only analyzed existing UK Biobank fine-mapping outputs, our V2D-MLP scores can directly be leveraged as priors in different fine-mapping contexts. When genotype-phenotype data are available and PolyFun ^11^ cannot be applied (e.g., the GWAS is not powerful enough to estimate trait-specific priors), V2D-MLP scores can be incorporated directly with SuSiE; when only GWAS summary statistics are available, they can also be incorporated into the ABF method ^38^. Finally, our V2D scores can be used to improve the power and accuracy of single-variant association tests ^39^ or polygenic risk scores ^40,41^. They could also be extended to rare variant burden tests ^42,43^, although further work is needed to assess how models trained on common and low-frequency variants generalize to rare variants.

We note several limitations of our work. First, the current V2D-MLP scores are limited to the annotations of the baseline-LF model and the 15 UK Biobank GWAS from European samples used for training. A critical next step is to improve V2D-MLP by incorporating more precise annotations (e.g., constraint scores, regulatory function ^13^, sequence context ^44^), and by leveraging *b*^2^ estimates from more independent traits and/or from multi-ancestry fine-mapping extensions of SuSiE ^45–47^. Second, we applied our V2D framework to effects averaged across 15 traits and did not investigate its application to a single GWAS for modeling trait-specific functional architectures. Indeed, applying the V2D framework to a single GWAS can be challenging because of potential limitations in statistical power and the availability of genome-wide fine-mapping results. However, our framework to compute trait-specific V2D-MLP scores offers a straightforward approach to overcoming these challenges. Third, the V2D-MLP framework can be computationally demanding: it requires estimating priors and performing genome-wide fine-mapping across large cohorts to generate training data and running computationally intensive machine learning models with grid search over millions of variants. Also, whether priors derived from the baseline-LF model must be updated when incorporating new annotations during training remains unclear. Despite these limitations, our results convincingly demonstrate the advantages of using our V2D framework to both characterize functional disease architectures and prioritize disease variants.

## Methods

### Applying the V2D framework to 15 UK Biobank traits

We applied the V2D framework to *b*^2^ estimated across 15 independent UK Biobank GWAS for ∼20M SNPs with MAF ≥ 0.1%. The 15 GWAS correspond to the 16 independent GWAS analyzed in ref. ^11^, from which we removed the trait hair color because of low polygenicity. For each trait, *b*^2^ estimates were obtained by PolyFun ^11^ using the default SuSiE with uniform priors (i.e., no functional priors; SuSiE-noprior) and with functional priors obtained from a linear combination of the annotations of the baseline-LF model as estimated by S-LDSC (SuSiE-prior). We assigned to each SNP *j* the mean value of normalized of *b*_j_^2^ (i.e., *b*_j_^2^/*Σ b* ^2^) across the 15 traits; within each trait, we removed SNPs explaining 1% of the total variance. We excluded SNPs from the MHC region (chr6:25-34Mb in hg19). *b*_j_^2^ were normalized so *Σb*_j_^2^ = 1.

We modeled *b*^2^ using the 187 annotations of the baseline-LF model ^3,7^ (**Table S2**). The baseline-LF model includes 10 MAF-bin annotations for common variants, 10 for low-frequency variants, 81 annotations each with both common and low-frequency counterparts (162 total), and 5 annotations for common variants only. Because analyses were run separately for common and low-frequency variants, we used 95 annotations for common variants and 90 for low-frequency variants, excluding the first MAF bin to avoid collinearity issues.

We evaluated four popular machine learning models: decision trees (python function `DecisionTreeRegressor`), MLP neural networks ^15^ (python function `MLPRegressor`), XGBoost ^16^ (python function `XGBRegressor`), and random forests ^17^ (python function `RandomForestRegressor`); a linear model (python function `LinearRegression`) was investigated as a baseline. The optimal set of hyperparameters was identified by a grid search minimizing MSE computed using a LEOCO cross-validation scheme. Specifically, for a given set of hyperparameters, we leveraged even (resp. odd) chromosomes to create a model, and odd (resp. even) chromosomes to compute the MSE; the total MSE was computed by weighting each MSE with the number of SNPs in even/odd chromosomes. This approach was applied using SuSiE- noprior *b*^2^ estimates, and selected hyperparameters were used for further analyses using SuSiE-noprior and SuSiE-prior estimates; we note that SuSiE-prior *b*_j_^2^ provided unreliable hyperparameters because the same set of chromosomes were used to compute priors and validate predictions. For decision trees, we varied the minimum number of leaf nodes (`min_samples_leaf` parameter) and the tree maximum depth (`max_depth`). For MLP, we varied the number of layers and neurons per layer (`hidden_layer_sizes`). For XGBoost, we varied the number of estimators (`n_estimators`), the maximum tree depth of base learner (`max_depth`), the minimum loss reduction for partition (`gamma`), the minimum sum of instance weight needed in a child (`min_child_weight`), and the subsample ratio of the training instances (`subsample`); we fixed learning rate (`learning_rate`) to 0.05. For random forests, we varied the minimum number of samples required to split an internal node (`min_samples_split`) and maximum depth of tree (`max_depth`); we fixed the number of base decision tree estimators (`n_estimators`) to 100. Because MLP convergence was sensitive to initialization, we ran each MLP with 10 random seeds. For LEOCO, we report mean validation MSE across seeds; for V2D score computation, we selected the seed with the highest training MSE. The full range of parameter values tested for each grid is available in **Table S12**. Grids included values both below and above the MSE minimum to ensure a thorough search for optimal parameters. All analyses were performed using standard packages in Python, with reproducible random seeds and default settings unless otherwise specified.

### Evaluating the V2D framework and V2D scores

We evaluated the V2D framework using heritability partitioning and excess overlap analyses on common variants. Analyses of prioritization scores were restricted to SNPs with a MAF > 5% in Europeans from the 1000 Genomes Project ^50^. V2D models were trained using common variants from UK Biobank, and we predicted V2D scores using annotations from the baseline-LD model v2.2 (we note that the baseline-LD v2.2 model contains the subset of common-variant annotations of the baseline-LF v2.2 model). To compare prioritization scores, we created annotations corresponding to the top X% of scores per chromosome; this design was motivated by the observation that some scores had uneven distributions of top variants across chromosomes and because our goal was to assess a score’s ability to prioritize variants at individual loci rather than genome-wide.

For common variants, heritability analyses were performed using S-LDSC (v1.0.1) with the baseline-LD model v2.2. We analyzed a set of 79 independent European GWAS constructed by removing from the 107 independent traits from ref. ^48^ the traits that were genetically correlated (*r_g_^2^*> 0.1 with ldsc ^49^) with the 15 UK Biobank GWAS. We also analyzed 20 independent East-Asian GWAS from ref. ^29^. For low-frequency variants, heritability analyses were performed using S-LDSC with the baseline-LF model v2.2 and following guidelines from ref. ^14^. We analyzed the set of 27 independent UK Biobank traits from ref. ^14^. We note that we were unable to construct a set of UK Biobank GWAS that were both genetically uncorrelated with the 15 GWAS and sufficiently powerful to evaluate low-frequency variant heritability explained by each leaf; however, in common variant analyses, we demonstrated the generalizability of the results obtained on the 15 GWAS to traits that are non-genetically correlated with them.

We evaluated prioritization scores on common variants by using excess overlap computed on 5 fine-mapping datasets (MVP ^13,19^, FinnGen ^20^, eQTLGen ^26,27^, Biobank Japan ^32^ and JCTF ^33^) and one MPRA dataset ^28^. Excess overlap was defined as the proportion of top scores within a set of positive variants divided by the proportion of top scores within a set of positive and negative variants. For each fine-mapping dataset, we define positive variants as those with a PIP > 0.90 in at least one trait/tissue, and the negative variants as those with a PIP < 0.01 in all traits/tissues (as performed in ref. ^13^). For the MPRA dataset, we define the positive variants as those that are defined as an expression modulating variant (emVar) in at least one cell line and the negative variants as those that have been tested but were not defined as an emVar in any cell line.

We evaluated six state-of-the-art prioritization scores: CADD ^21^, Eigen ^22^, GWAVA ^23^, LINSIGHT ^24^, ReMM ^25^ and cV2F ^13^. For GWAVA, we report results for region scores because they provided higher enrichment and excess overlap than unmatched scores and TSS scores. We note that cV2F scores are trained on 339 features, including 84 annotations from the baseline-LD model and 12 annotations constructed from the MPRA dataset used for validation. For comparability, we therefore 1) recomputed cV2F scores using the same annotations as V2D-MLP (we labeled these scores cV2F-bLD) for comparison purposes and 2) evaluated cV2F-bLD, rather than cV2F, on the MPRA dataset to avoid circularity.

### Simulations to evaluate SuSiE *b^2^* estimates

We simulated GWAS summary statistics from 541,126 common SNPs of chromosome 1, an LD matrix estimated on 337,491 unrelated British-ancestry individuals from UK Biobank release 3 (ref.^11^), 0.5% of causal SNPs, a sample size of *N* = 350K (approximating the LD matrix sample size and the GWAS maximum sample size analyzed in this study). We simulated under different functional architectures (defined with E(*β*_j_) = 0 and different Var(*β*_j_); see next paragraph) and different values of *h*^2^ (0.5, 0.2, and 0.1). First, causal SNPs were sampled with probability proportional to Var(*β*_j_). Second, effect sizes were drawn from a normal distribution, and normalized the effects to the value set for *h*^2^ (i.e., *Σβ*_j_^2^ = *h*^2^). Third, GWAS Z-scores were simulated using a multivariate normal distribution (*mvrnorm R* function) and 78 non-overlapping 3Mb UK Biobank LD matrices.

Finally, SuSiE was run on each LD block using UK Biobank LD matrices; for SuSiE-prior, priors were estimated on an independent simulation with similar parameters (i.e., functional architectures and *h*^2^). For each simulation scenario, we simulated 100 summary statistics. We evaluated SuSiE-noprior and SuSiE-prior *b*^2^ estimates by comparing the true *h*^2^ functional enrichment of annotations obtained from simulated *β*^2^ (i.e., the mean *β*_j_^2^ for all variants within an annotation divided by the mean *β*_j_^2^ for all variants) to those obtained from estimated *b*^2^.

We considered two functional architectures. In the first architecture, we defined Var(*β*_j_) using a linear combination of functional annotations. Specifically, we used the S-LDSC model, which assumes that Var(*β*_j_) is proportional to *Σ*_k_*τ*_k_*a*_k,j_ where *τ*_k_ is the effect of annotation *k* on *h*^2^ and *a*_k,j_ is the value of annotation *k* for variant *j*; here, we used the 97 annotations from the baseline-LD model v2.2, and *τ*_k_ obtained by averaging their S-LDSC estimates across the 15 independent UK Biobank traits. We set negative values of Var(*β*_j_) to 0. For SuSiE-prior, priors were estimated by regressing effects on all the annotations of the baseline-LD model. In the second architecture, we defined Var(*β*_j_) using a non-linear combination of functional annotations to investigate scenarios in which the linear model used by SuSiE-prior is not matching the generative model. Specifically, we considered a simple model depending only on the coding and conserved annotations, and their interaction, and defined Var(*β*_j_) proportional to 1 + coding_j_ + 5 x conserved_j_ + 11 x coding_j_ x conserved_j_, where coding_j_ (resp. conserved_j_) is an indicator function taking the value 1 if variant belongs to the coding (resp. conserved) annotation. Coefficients were manually selected to yield realistic *h*^2^ enrichments for coding and conserved annotations. For SuSiE-prior, priors were estimated by regressing effects only on the coding and conserved annotations. S-LDSC was run using only the coding and conserved annotations (as performed in practice) and also by adding an annotation corresponding to their interaction (S-LDSC+interaction).

### V2D-informed fine-mapping of 110 UK Biobank traits

We performed functionally-informed fine-mapping of 110 UK Biobank traits using the approach from refs. ^12,13^. Briefly, this method reweights posterior probabilities of variants within SuSiE credible sets (CS) by using external prioritization scores. Here, we leveraged the V2D-MLP model trained on common variants and computed V2D-MLP scores for all variants with available cV2F scores (i.e., 10 million variants with a minor allele count ≥ 5 in Europeans from the 1000 Genomes Project ^50^) to allow one-by-one comparison. For V2D- MLP informed fine-mapping, we directly used V2D-MLP scores to reweight posterior probabilities. For cV2F informed fine-mapping, cV2F scores lower than 0.75 were converted to 0.1, and scores higher than 0.75 were converted to 1 (as performed in ref. ^13^). To compare two fine-mapping methods, we focused on CS in which the top-PIP variant differed between the two approaches and both variants had been assayed by MPRA. We then compared the proportion of emVars using McNemar’s paired test.

We computed trait-specific V2D-MLP scores proportional to *τ*_0_ + *τ*_1_ V2D + Σ_a_ [*τ*_a_ CTS_a_ + *τ*_a*_ CTS_a_ x V2D], where V2D is the vector of V2D-MLP scores, *a* is the number of cell-type-specific annotations to include, CTS_a_ is the vector of cell-type-specific annotation *a*, and *τ*_x_ are linear coefficients; we identified the CTS annotations to include using CT-FM ^34^ and estimated *τ*_x_ using S-LDSC with a LEOCO scheme from the trait summary statistics. We created trait-specific V2D scores for seven blood-related UK Biobank traits that had both SuSiE and CT-FM results available and that were well-powered for functionally-informed fine-mapping: mean corpuscular hemoglobin level (279 SNPs with PIP > 0.5 with SuSiE), monocyte count (239 SNPs), red blood cell count (219 SNPs), eosinophil count (212 SNPs), lymphocyte count (189 SNPs), white blood cell count (174 SNPs), and asthma (28 SNPs).

## Data availability

V2D-MLP scores, posterior mean squared causal effect sizes of 15 UK Biobank traits, and annotations of the baseline-LF model version 2.2: https://zenodo.org/records/17257765.

S-LDSC reference files and GWAS summary statistics: https://zenodo.org/records/10515792 and https://zenodo.org/records/7787039.

East-Asian GWAS summary statistics: https://zenodo.org/records/11455096.

MVP fine-mapping dataset: https://mskcc.ent.box.com/s/9gn1h0etn8efo8hlnven3qz1qufupm6m/. FinnGen fine-mapping dataset: https://elomake.helsinki.fi/lomakkeet/124935/lomake.html.

UK Biobank fine-mapping results: https://www.finucanelab.org/data and gs://finucane-requester-pays/ukbb-finemapping/.

GTEx fine-mapping dataset: https://www.finucanelab.org/data.

eQTLGen fine-mapping dataset: https://zenodo.org/records/10117202 (file S2G_original.zip). Biobank Japan fine-mapping dataset: https://pheweb.jp/downloads.

JCTF fine-mapping dataset: https://humandbs.dbcls.jp/files/hum0343/hum0343.v3.qtl.v1.zip. MPRA dataset: Supplementary Table 3 of ref. ^28^.

CADD (version 1.7) scores: https://cadd.gs.washington.edu/download. Eigen scores: https://zenodo.org/communities/iuliana-ionita-laza/.

GWAVA scores: ftp://ftp.sanger.ac.uk/pub/resources/software/gwava/v1.0/annotated/gwava_db_csv.tgz. LINSIGHT scores: http://compgen.cshl.edu/LINSIGHT/.

ReMM scores: https://remm.bihealth.org/download.

cV2F scores: https://mskcc.box.com/shared/static/hsrogtr3fddtmd53hyy5ph7dlp20eq72.txt.

## Code availability

The software and the code to replicate our analyses are available at https://github.com/chengsly/V2D.

## Data Availability

Data availability
V2D-MLP scores, posterior mean squared causal effect sizes of 15 UK Biobank traits, and annotations of the baseline-LF model version 2.2: https://zenodo.org/records/17257765.
S-LDSC reference files and GWAS summary statistics: https://zenodo.org/records/10515792 and https://zenodo.org/records/7787039.
East-Asian GWAS summary statistics: https://zenodo.org/records/11455096.
MVP fine-mapping dataset: https://mskcc.ent.box.com/s/9gn1h0etn8efo8hlnven3qz1qufupm6m/.
FinnGen fine-mapping dataset: https://elomake.helsinki.fi/lomakkeet/124935/lomake.html.
UK Biobank fine-mapping results: https://www.finucanelab.org/data and gs://finucane-requester-pays/ukbb-finemapping/.
GTEx fine-mapping dataset: https://www.finucanelab.org/data.
eQTLGen fine-mapping dataset: https://zenodo.org/records/10117202 (file S2G_original.zip).
Biobank Japan fine-mapping dataset: https://pheweb.jp/downloads.
JCTF fine-mapping dataset: https://humandbs.dbcls.jp/files/hum0343/hum0343.v3.qtl.v1.zip.
MPRA dataset: Supplementary Table 3 of ref. 28.
CADD (version 1.7) scores: https://cadd.gs.washington.edu/download.
Eigen scores: https://zenodo.org/communities/iuliana-ionita-laza/.
GWAVA scores: ftp://ftp.sanger.ac.uk/pub/resources/software/gwava/v1.0/annotated/gwava_db_csv.tgz.
LINSIGHT scores: http://compgen.cshl.edu/LINSIGHT/.
ReMM scores: https://remm.bihealth.org/download.
cV2F scores: https://mskcc.box.com/shared/static/hsrogtr3fddtmd53hyy5ph7dlp20eq72.txt.

## Acknowledgments

We thank N. Mancuso, members of the Gazal and Mancuso labs, and K. Dey for helpful discussions. We thank the International Multiple Sclerosis Genetics Consortium (IMSGC) for sharing multiple sclerosis GWAS summary statistics. This research has been funded by the National Institutes of Health grant R35 GM147789.

## Supplementary Figures

**Figure S1.**
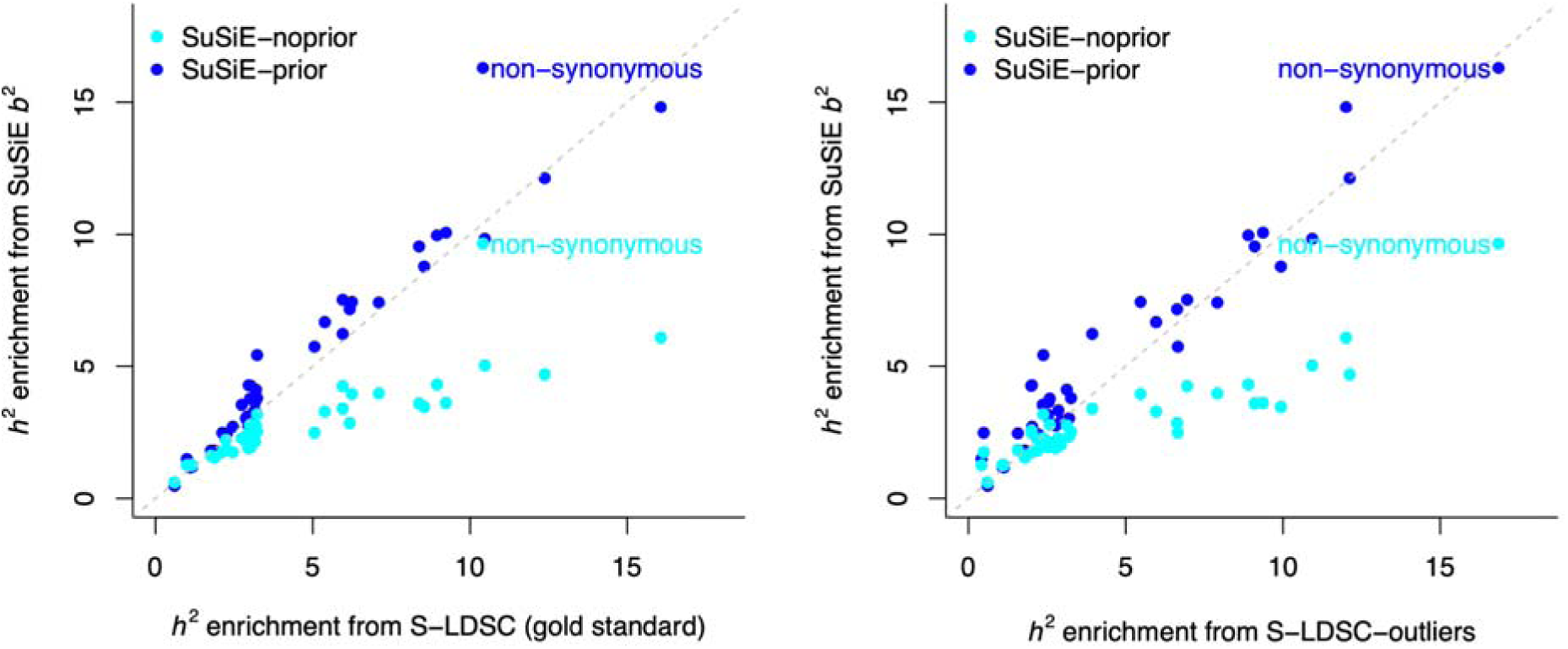
Comparing SuSiE *b*^2^ estimates against S-LDSC for 15 UK Biobank traits. We report heritability (*h*^2^) enrichment estimated using S-LDSC, SuSiE-noprior, and SuSiE-prior (priors obtained with PolyFun). The left panel is similar to Fig. 3A and shows default S-LDSC outputs, which exclude variants with high ch -square statistics. The right panel shows S-LDSC outputs obtained when including all the variants (S-LDSC-outliers); values were obtained by using the ldsc `--chisq-max 9999` option. For the non-synonymous annotation, S-LDSC-outliers yield higher enrichment than default S-LDSC, which suggests that default filtering underestimates *h^2^* enrichment for this annotation.

**Figure S2.**
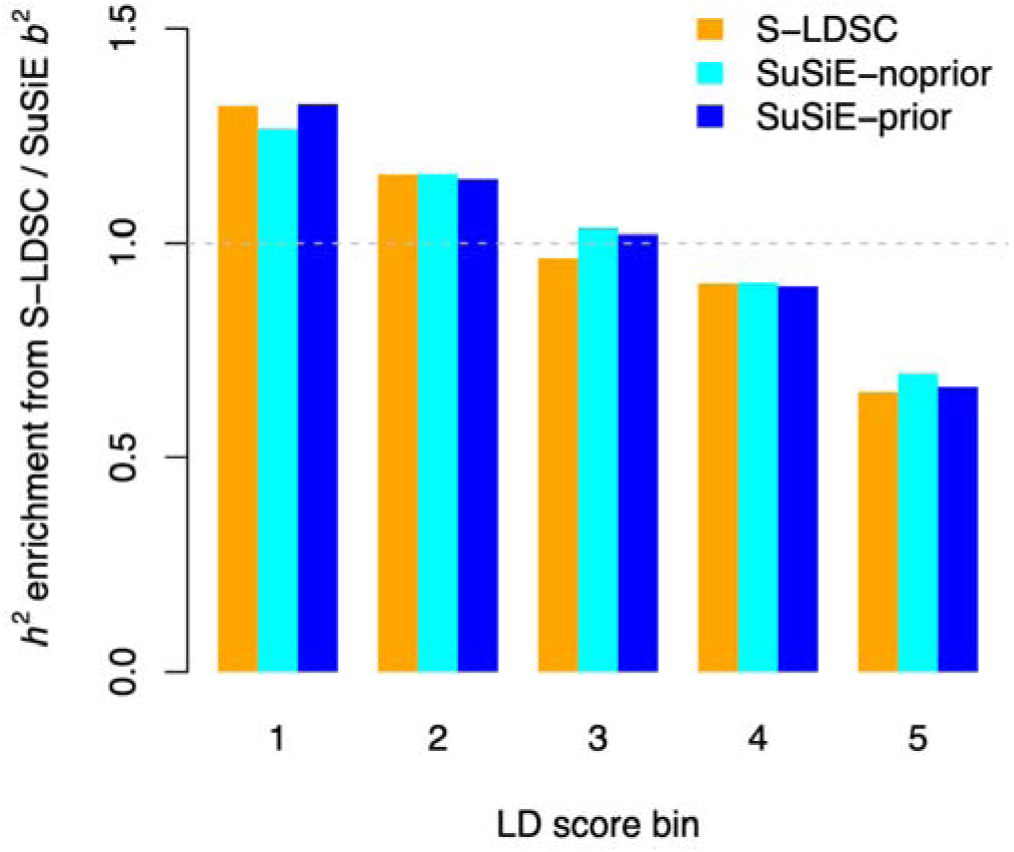
Evaluating SuSiE *b*^2^ estimates on 15 UK Biobank traits. We report heritability enrichment estimated with S-LDSC (gold standard), SuSiE-noprior, and SuSiE-prior (PolyFun priors) across quintiles of LD scores. Numerical results and standard errors are reported in **Table S7**.

**Figure S3.**
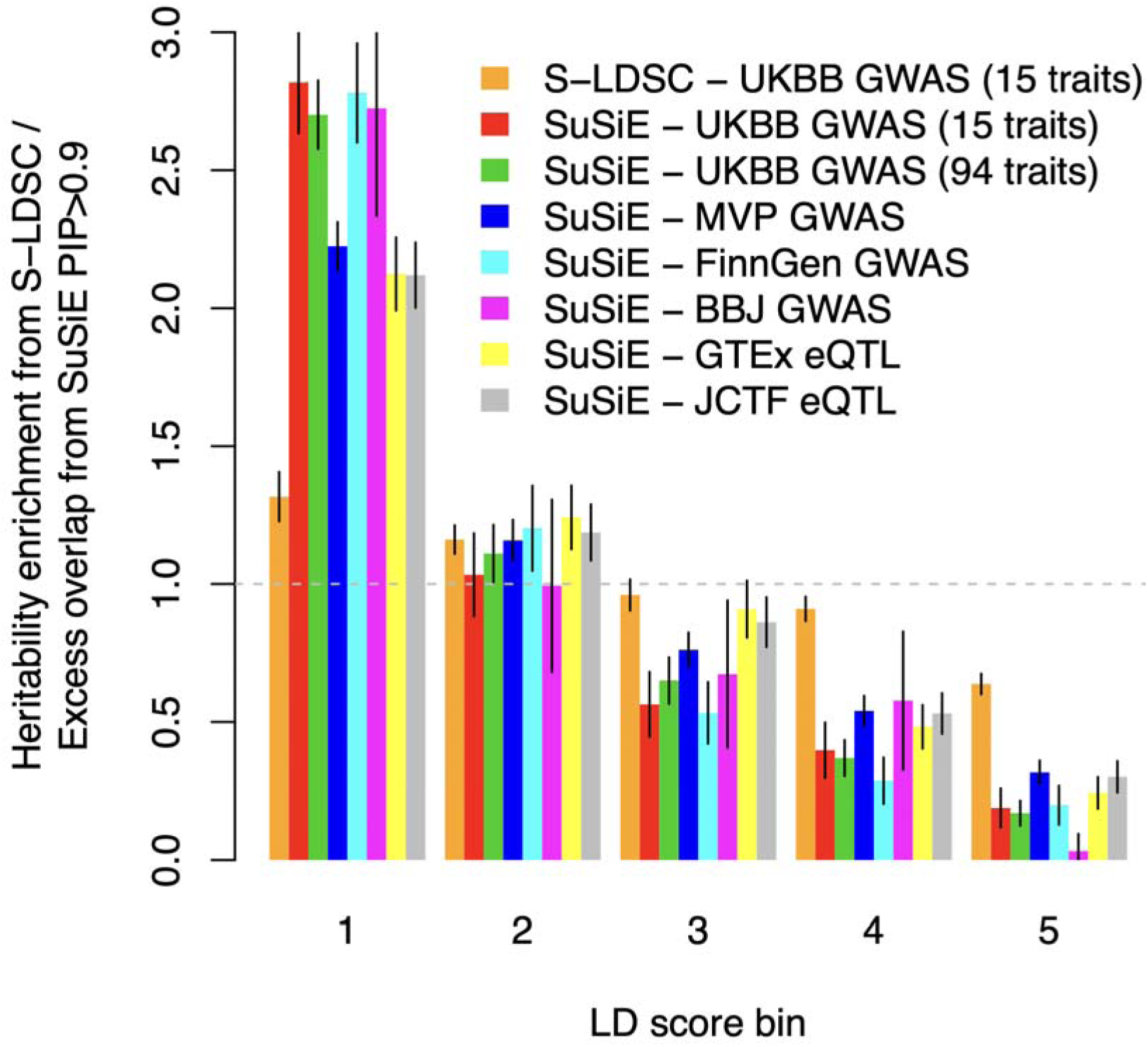
Variants that are confidently fine-mapped tend to have low LD scores. We report the excess overlap of variants that were confidently fine-mapped with SuSiE (PIP > 0.9) across seven datasets: the 15 independent UK Biobank traits used in this study (from ref. ^11^), the 94 UK Biobank traits used in ref. ^13^, the 931 Million Veteran Program (MVP) GWAS used in refs. ^13,19^, FinnGen ^20^, Biobank Japan (BBJ) ^32^, eQTLs from GTEx ^51,52^, and the Japan COVID-19 Task Force (JCTF) ^33^. S-LDSC estimates averaged across the 15 independent UK Biobank traits as a gold standard (here in orange) because we demonstrated that S-LDSC estimates for LD-related annotations are unbiased in simulations ^3^. Variants fine-mapped with high confidence were disproportionately enriched in the lowest LD bin, which is consistent with their greater ease of fine-mapping.

**Figure S4.**
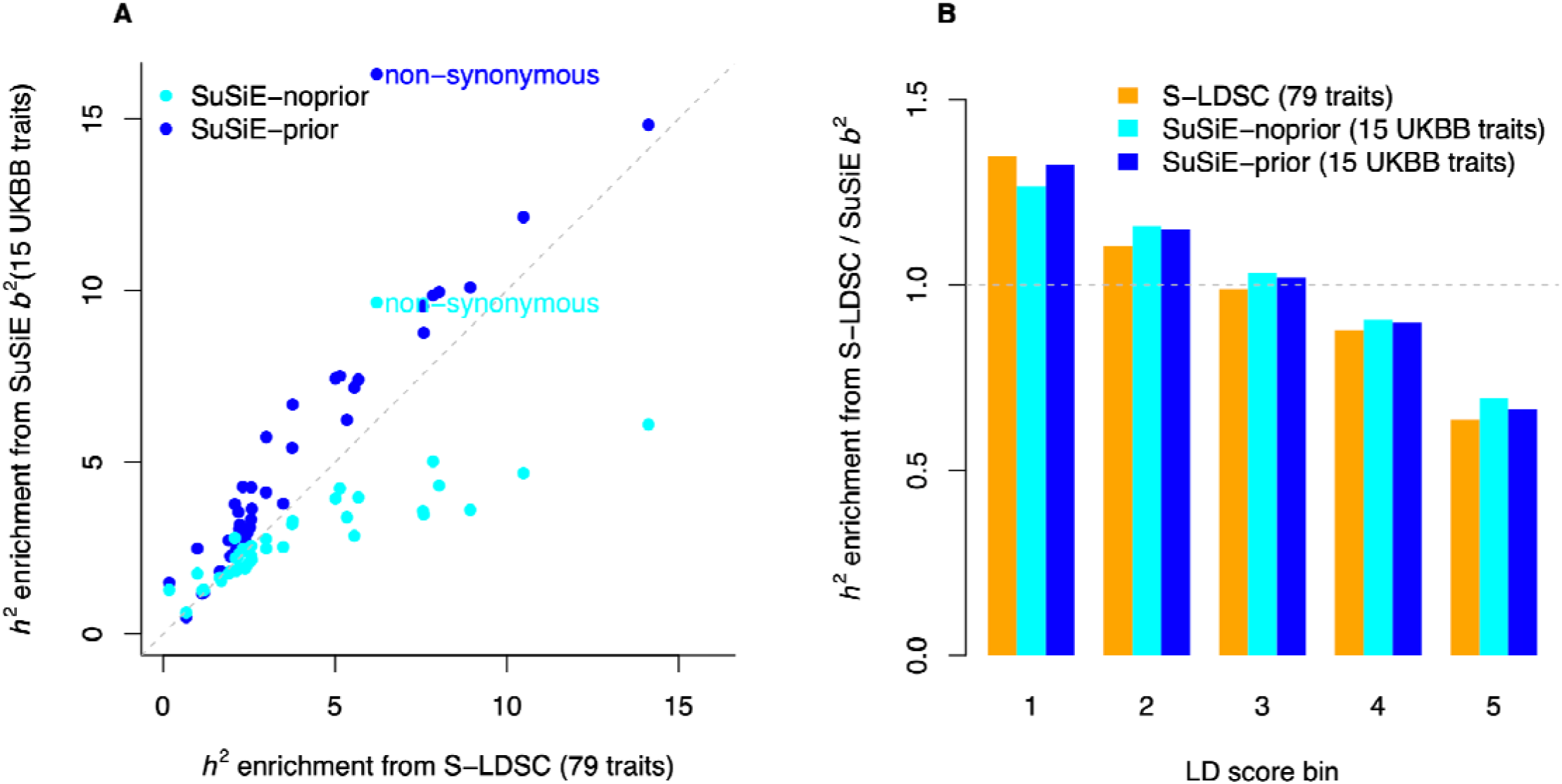
Comparing SuSiE *b*^2^ estimates on 15 UK Biobank traits against S-LDSC on 79 independent GWAS. (**A**) We report heritability enrichment for 40 main functional annotations, comparing SuSiE-noprior and SuSiE-prior (PolyFun priors) from 15 UK Biobank traits with S-LDSC estimates from 79 independent GWAS. **(B)** We report heritability enrichment within quintiles of LD scores. Results were similar to those in Fig. 3, indicating that SuSiE _j_^2^ estimates from 15 UK Biobank traits are representative of human diseases and complex trait genetic architecture.

**Figure S5.**
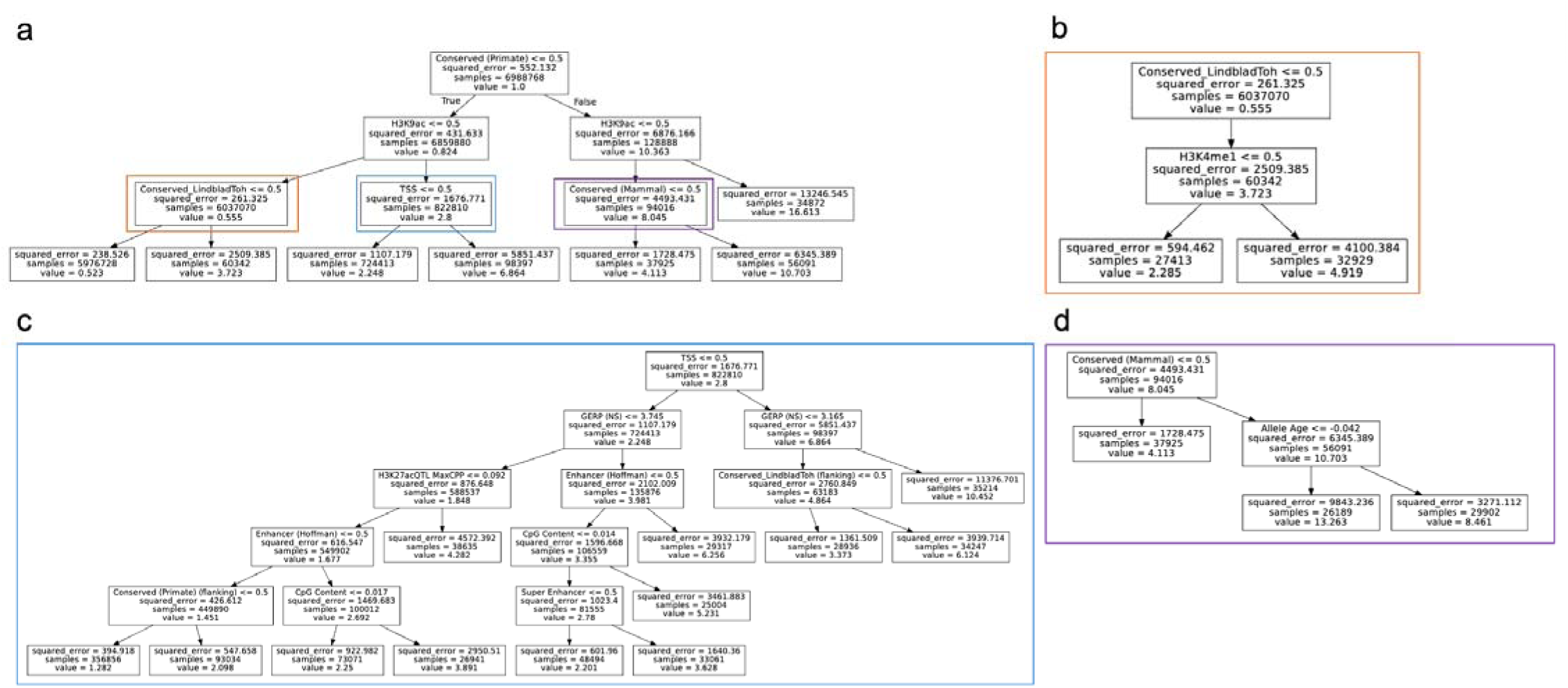
Decision tree using SuSiE-prior _j_^2^ on common variants. We report a decision tree with depth = 7 and minimum number of common SNPs per leaf = 25K obtained on normalized SuSiE-prior _j_^2^ estimates averaged across 15 UK Biobank traits. **(a)** Root of the tree (depth = 3). **(b–d)** Deeper branches. Each leaf reports the mean effect of the variant (value; normalized so the average effect of all common variants = 1), the number of SNPs in the leaf (samples), and the mean squared error (squared_error).

**Figure S6.**
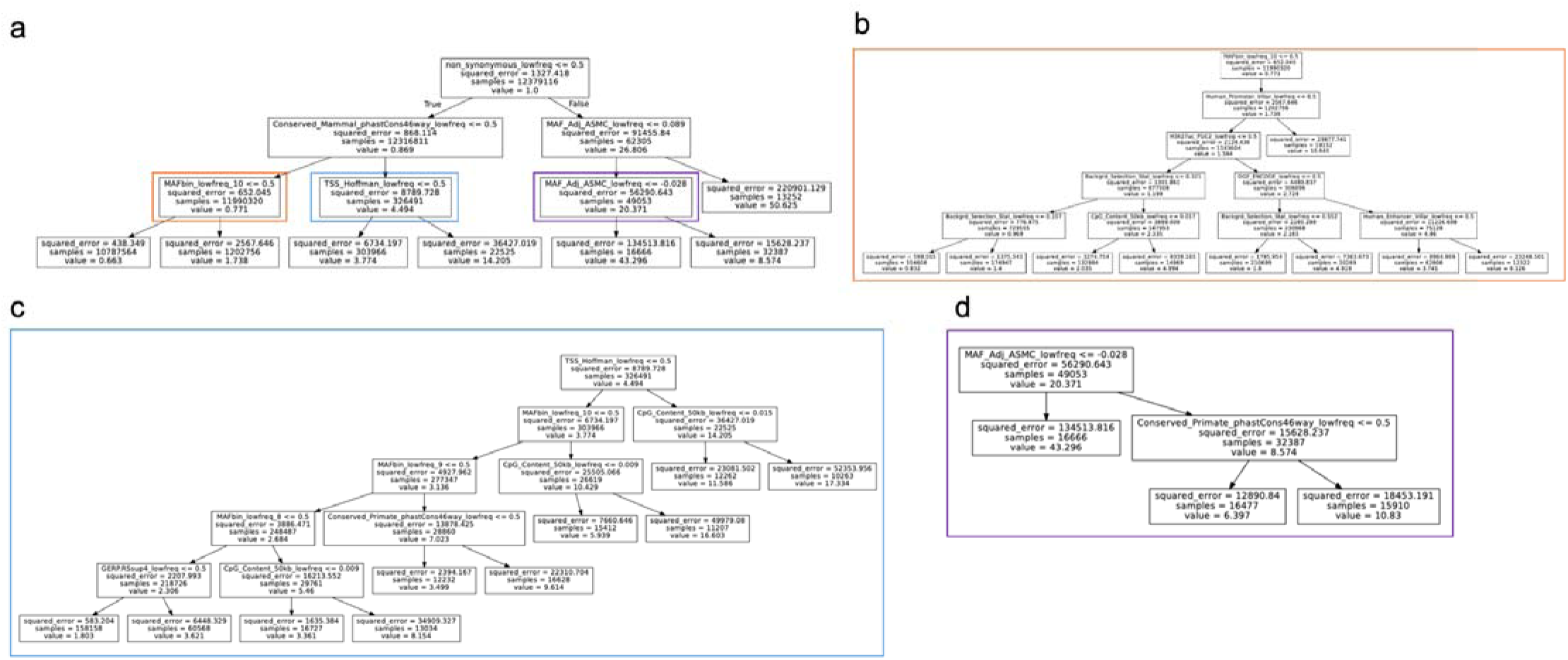
Decision tree using SuSiE-prior _j_^2^ on low-frequency variants. We report a decision tree with depth = 7 and minimum number of low-frequency SNPs per leaf = 10K obtained on normalized SuSiE-prior _j_^2^ estimates averaged across 15 UK Biobank traits. **(a)** Root of the tree (depth = 3). **(b–d)** Deeper branches. Each leaf reports the mean effect of the variant (value; normalized so the average effect of all low-frequency variants = 1), the number of SNPs in the leaf (samples), and the mean squared error (squared_error).

**Figure S7.**
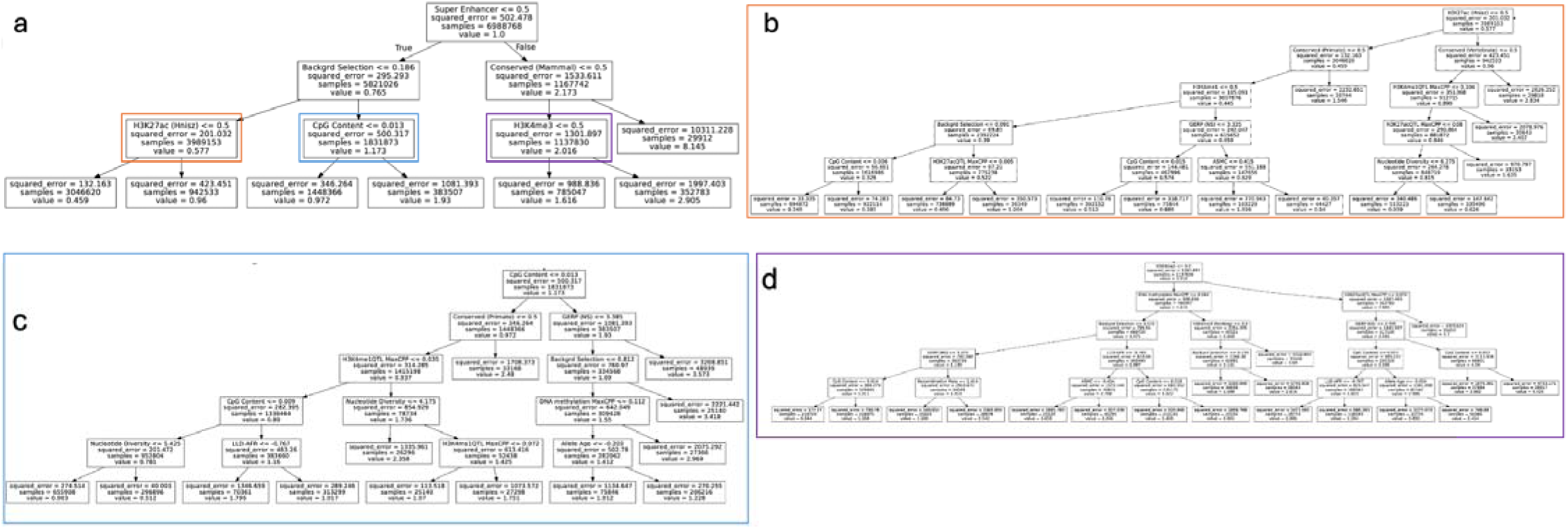
Decision tree using SuSiE-noprior _j_^2^ on common variants. We report a decision tree with depth = 7 and minimum number of common SNPs per leaf = 25K obtained on normalized SuSiE-noprior _j_^2^ estimates averaged across 15 UK Biobank traits. **(a)** Root of the tree (depth = 3). **(b–d)** Deeper branches. Each leaf reports the mean effect of the variant (value; normalized so the average effect of all common variants = 1), the number of SNPs in the leaf (samples), and the mean squared error (squared_error).

**Figure S8.**
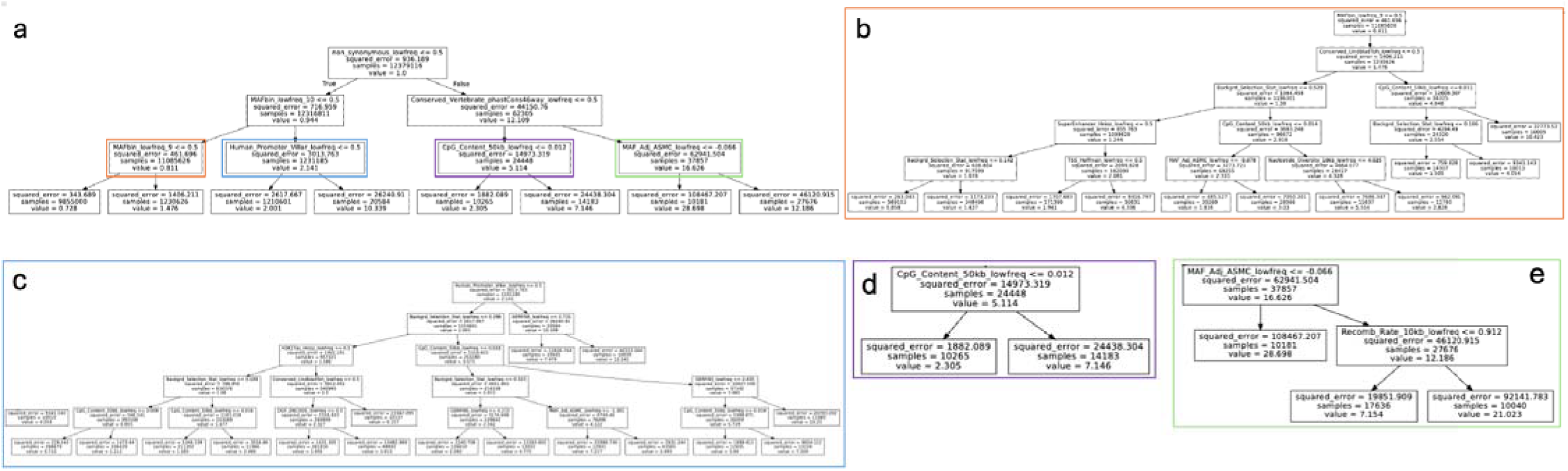
Decision tree using SuSiE-noprior _j_^2^ on low-frequency variants. We report a decision tree with depth = 7 and minimum number of low-frequency SNPs per leaf = 10K obtained on normalized SuSiE-noprior _j_^2^ estimates averaged across 15 UK Biobank traits. **(a)** Root of the tree (depth = 3). **(b–d)** Deeper branches. Each leaf reports the mean effect of the variant (value; normalized so the average effect of all low-frequency variants = 1), the number of SNPs in the leaf (samples), and the mean squared error (squared_error).

**Figure S9.**
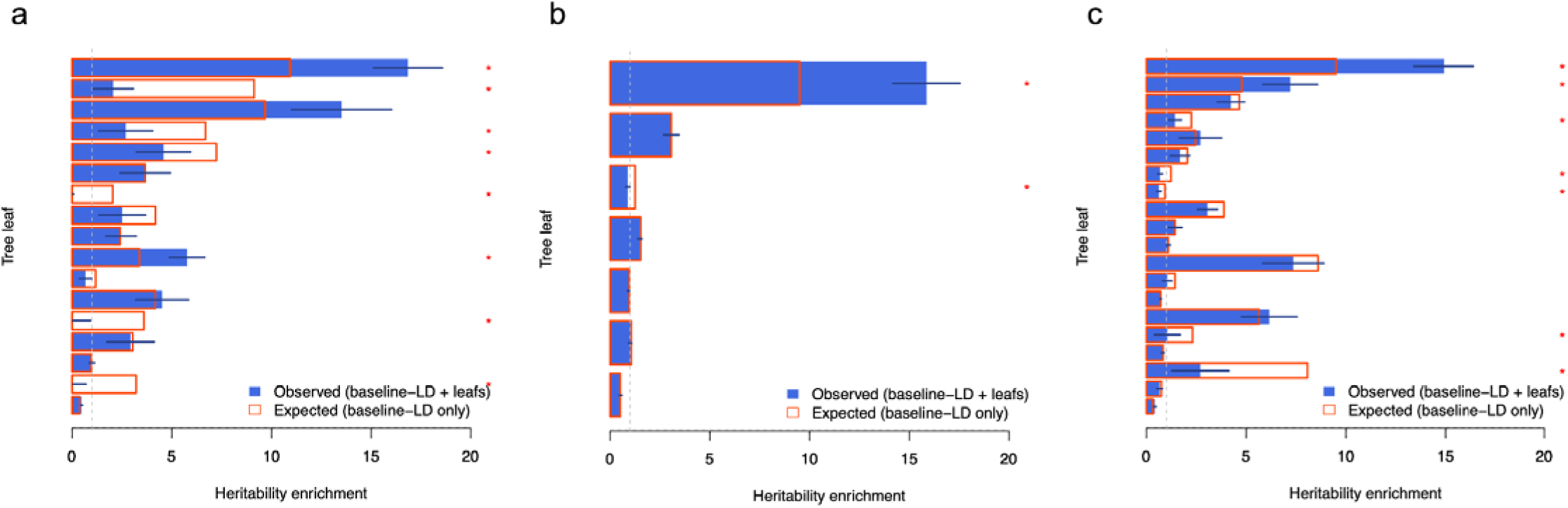
Heritability enrichment of common SNPs in leaves from decision trees. We report heritability enrichment of each leaf estimated with S-LDSC (blue) and compared to expectations under the baseline-LD model (red) using 79 independent European GWAS not overlapping the 15 UK Biobank traits. Error bars represent 95% confidence intervals, and asterisks indicate significant differences after Bonferroni correction. **(a)** We report enrichments for leaves constructed using SuSiE-prior _j_^2^ on decision tree with depth = 5; the leaf order (from bottom to top) is the same as plotted in **Fig. S5** (i.e., the top leaf corresponds to constrained in primates and in H3K9ac peaks). **(b)** We report enrichments for leaves constructed using SuSiE-noprior _j_^2^ on decision tree with depth = 3; the leaf order (from bottom to top) is the same as plotted in **Fig. S7**. **(c)** We report enrichments for leaves constructed using SuSiE-noprior _j_^2^ on a decision tree with depth = 5; the leaf order (from bottom to top) is the same as plotted in **Fig. S7**.

**Figure S10.**
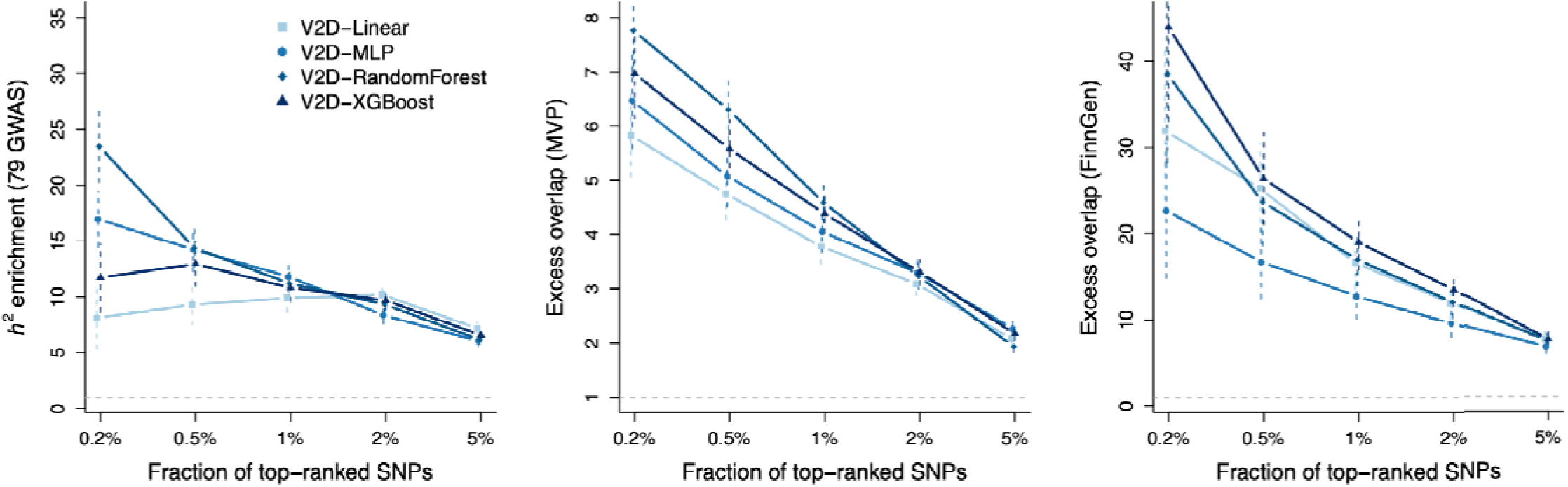
V2D scores obtained when using SuSiE-noprior _j_^2^. We report heritability enrichment across 79 independent GWAS and excess overlap with fine-mapped variants from MVP and FinnGen computed for V2D scores derived from linear, MLP, random forest, and XGBoost models. Error bars represent 95% confidence intervals.

**Figure S11.**
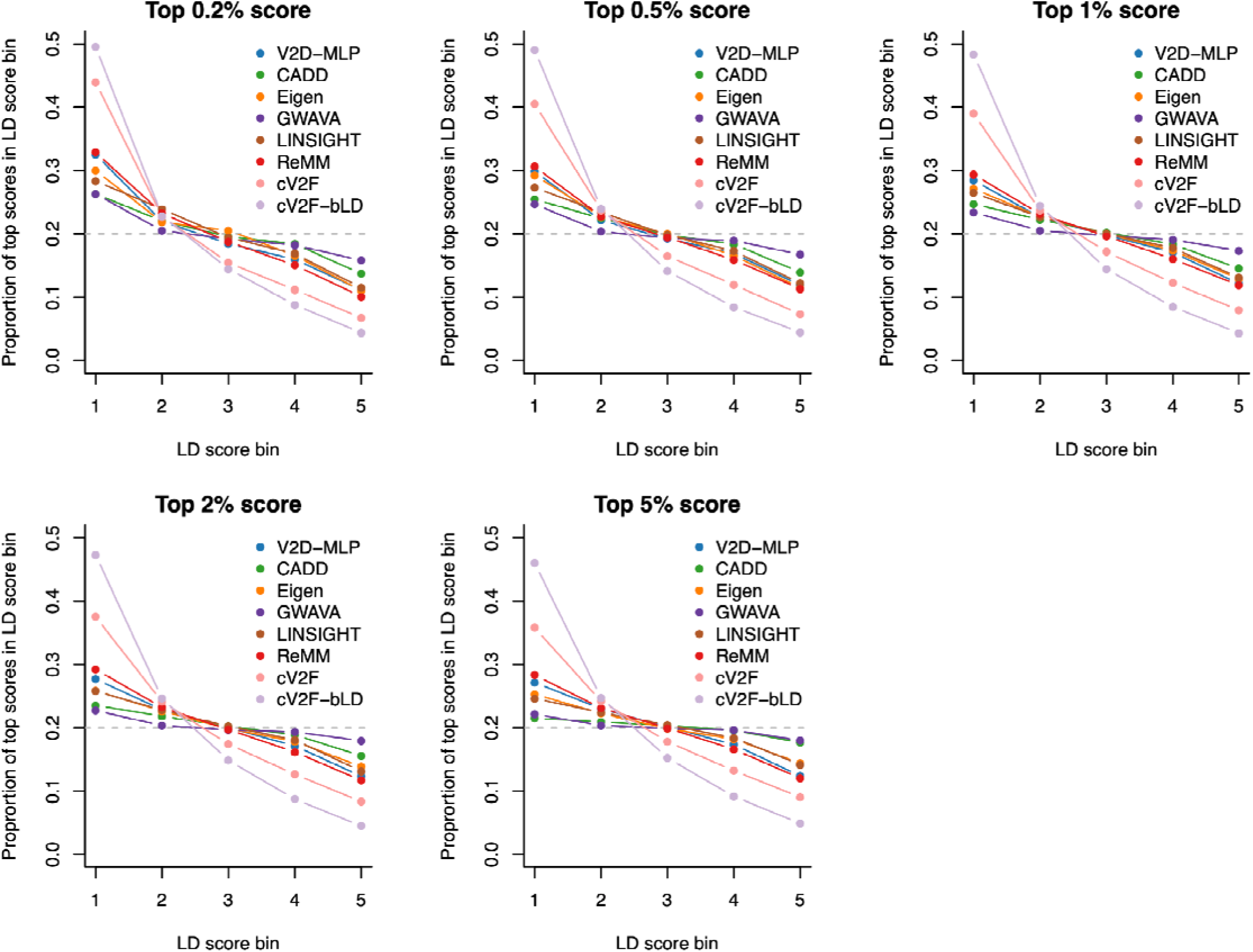
Proportion of top prioritization scores in LD score quintiles. We report the proportion of top-ranked variants within each LD score quintile for multiple prioritization scores. cV2F and cV2F-bLD are highly enriched (∼2×) in low-LD variants but markedly depleted (∼0.5×) in high-LD variants, thus reflecting their training on fine-mapped variants with high precision, which tend to occur in regions of low LD (see **Fig. S3**).

**Figure S12.**
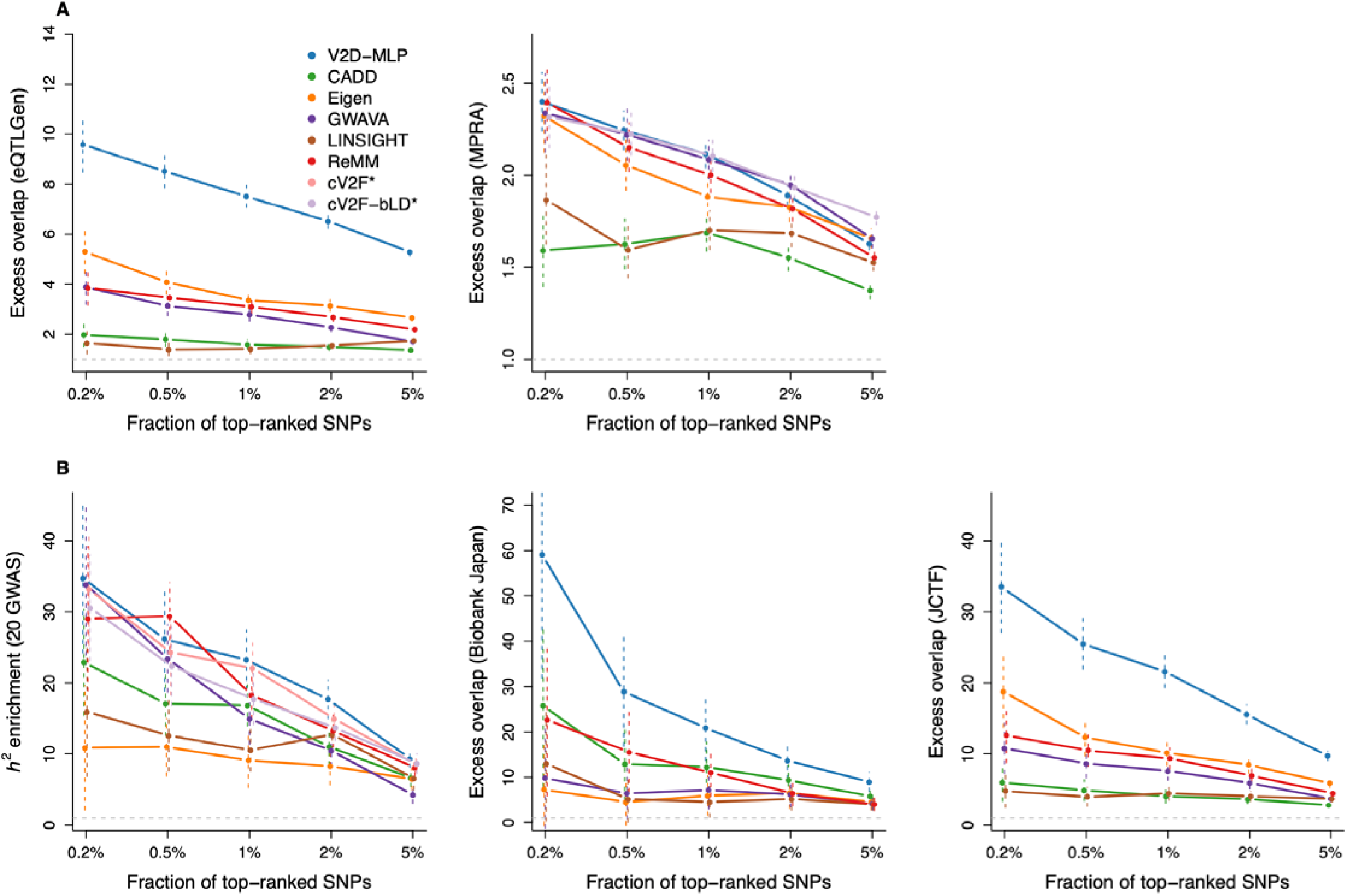
Benchmarking V2D scores across gene expression phenotypes and non-European ancestry datasets. **(A)** We report excess overlap on fine-mapped eQTLs from eQTLGen as well as 233K variants functionally tested using massively parallel reporter assay (MPRA) across five cell lines. **(B)** We report *h^2^*enrichment computed on 20 independent East-Asian GWAS and excess overlap computed on fine-mapped variants from 79 Biobank Japan GWAS and eQTLs from the Japan COVID-19 Task Force (JCTF). * cV2F and cV2F-bLD were not evaluated using fine-mapped datasets because cV2F scores are specifically trained on variants fine-mapped with high confidence and are inherently enriched toward SNPs with low LD versus other prioritization scores (**Figs. S3** and **S11**); cV2F was also not evaluated with the MPRA dataset because the same MPRA dataset is used as a feature of cV2F model.

**Figure S13.**
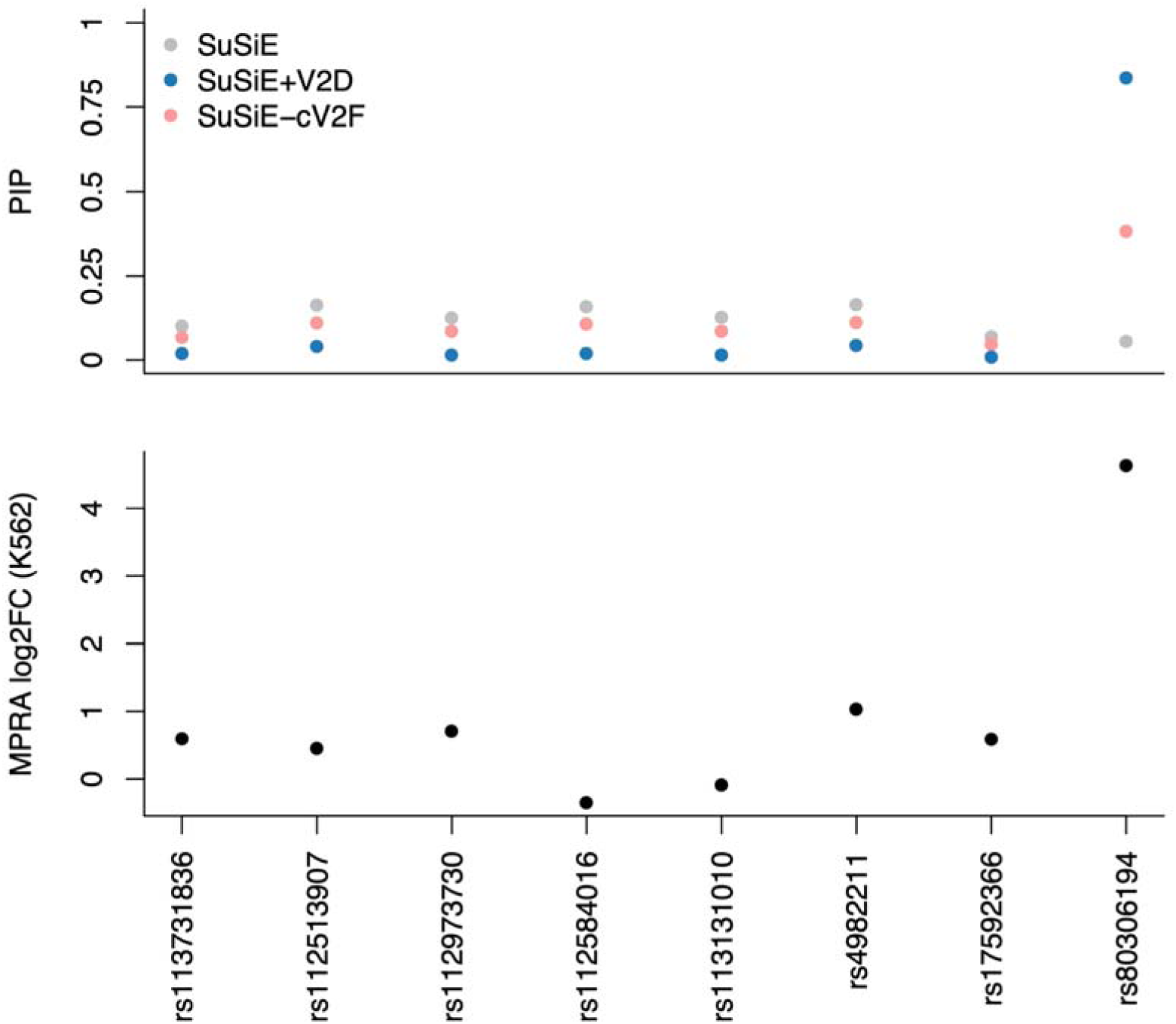
Fine-mapping and MPRA results at the *BAZ1A* locus of the red blood cell count GWAS. We report the posterior inclusion probabilities (PIPs) from SuSiE, SuSiE+V2D, and SuSiE+cV2F for the eight SNPs in the credible set (top). Corresponding MPRA log2 fold-change (log2FC) values in K562 cells are shown below (bottom).

**Figure S14.**
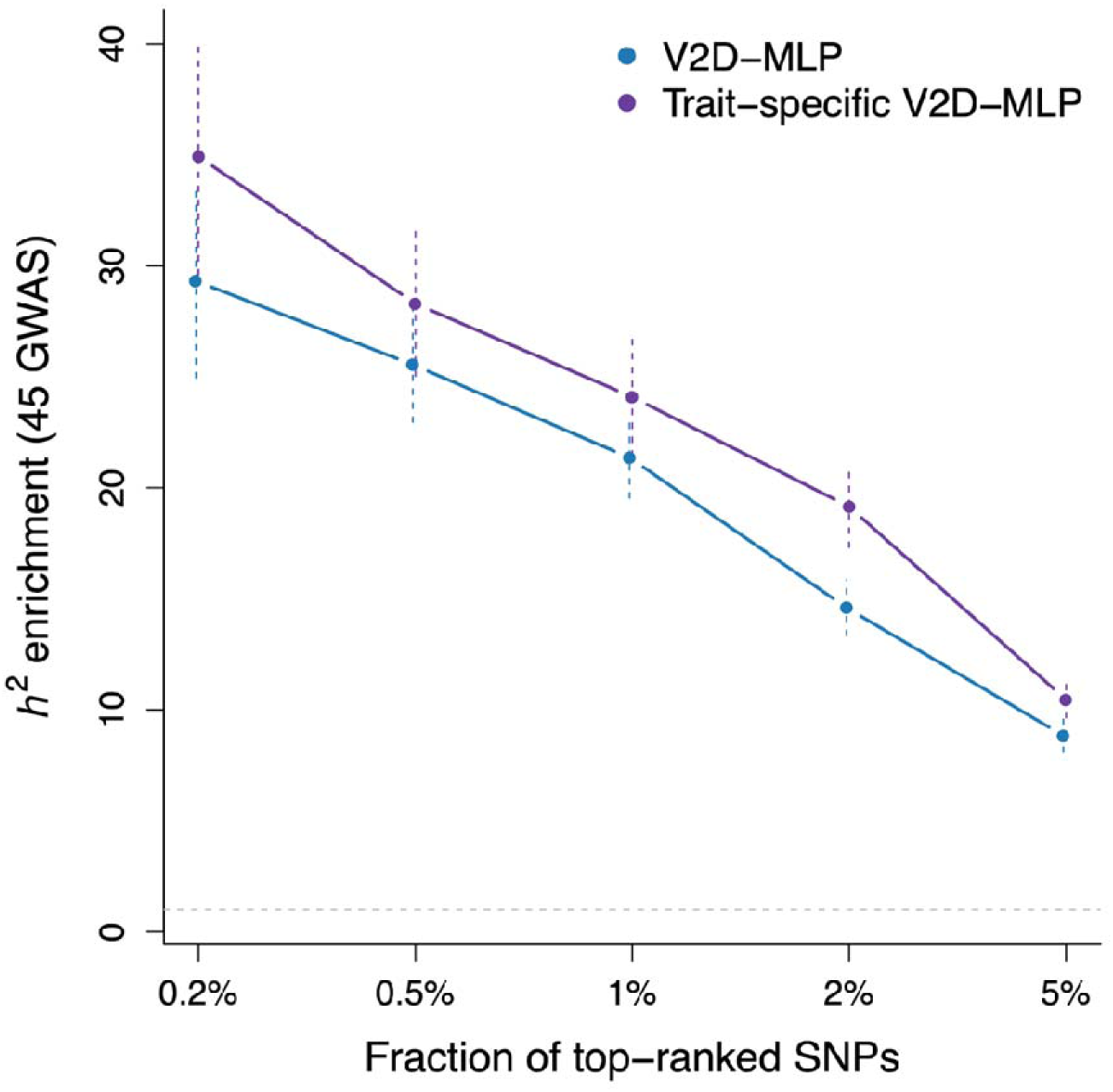
Heritability enrichments for V2D-MLP and trait-specific V2D-MLP scores. We report *h^2^* enrichment computed across 45 of 79 independent GWAS for which we were able to create trait-specific V2D-MLP scores. Error bars represent 95% confidence intervals. Enrichment obtained with trait-specific V2D-MLP scores were consistently higher than those obtained with V2D-MLP scores; differences were significant for the top 2% SNPs (*P* = 5.7 x 10^-5^) and top 5% SNPs (*P* = 0.004); we note that the cell-type-specific annotations leveraged to compute trait-specific V2D-MLP scores captured a mean of 1.5% of common SNPs.

**Figure S15.**
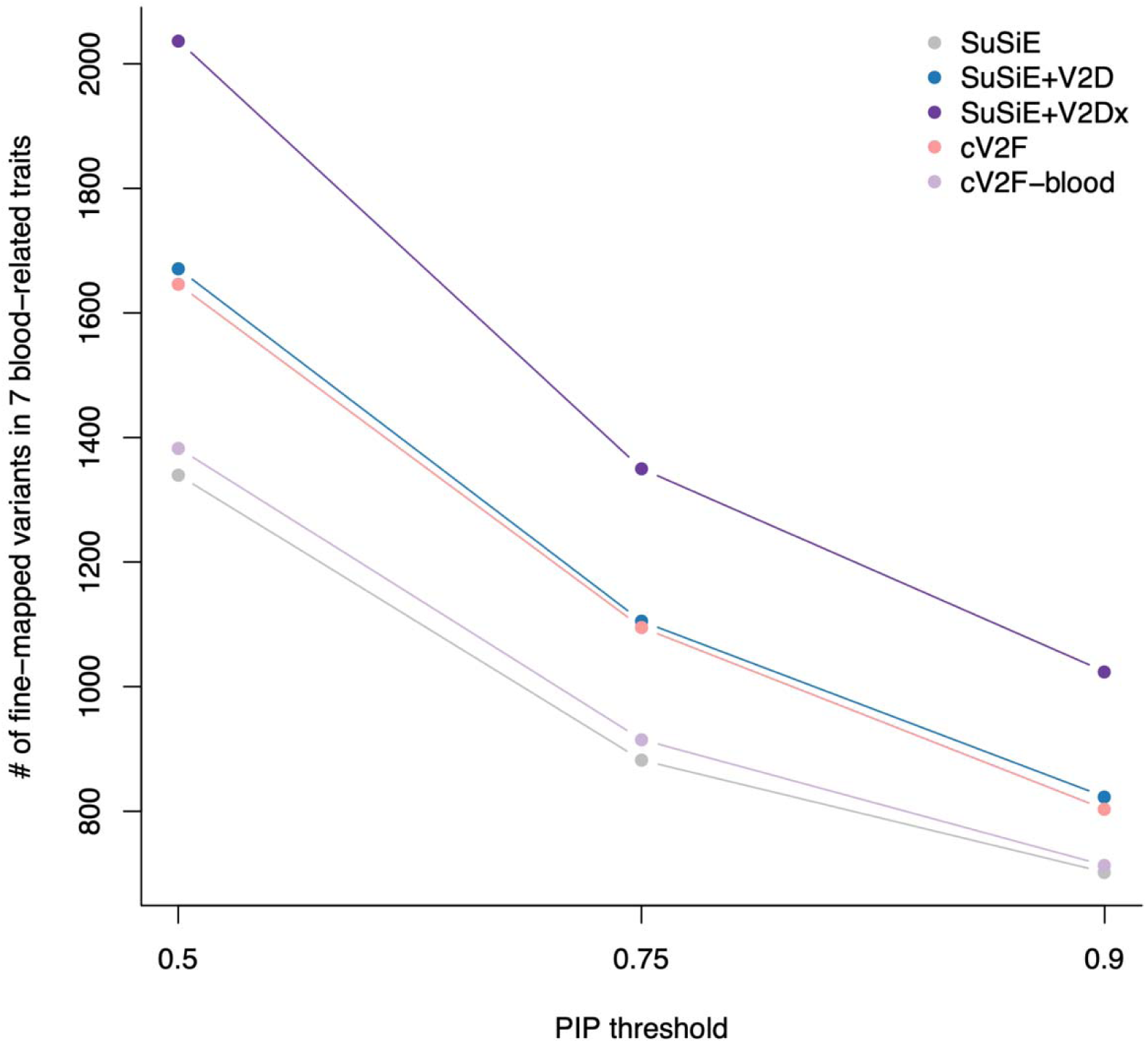
Leveraging trait-specific V2D scores to prioritize variants of blood-related traits. We report the number of fine-mapped SNPs exceeding PIP thresholds across seven UK Biobank blood-related traits.

**Figure S16.**
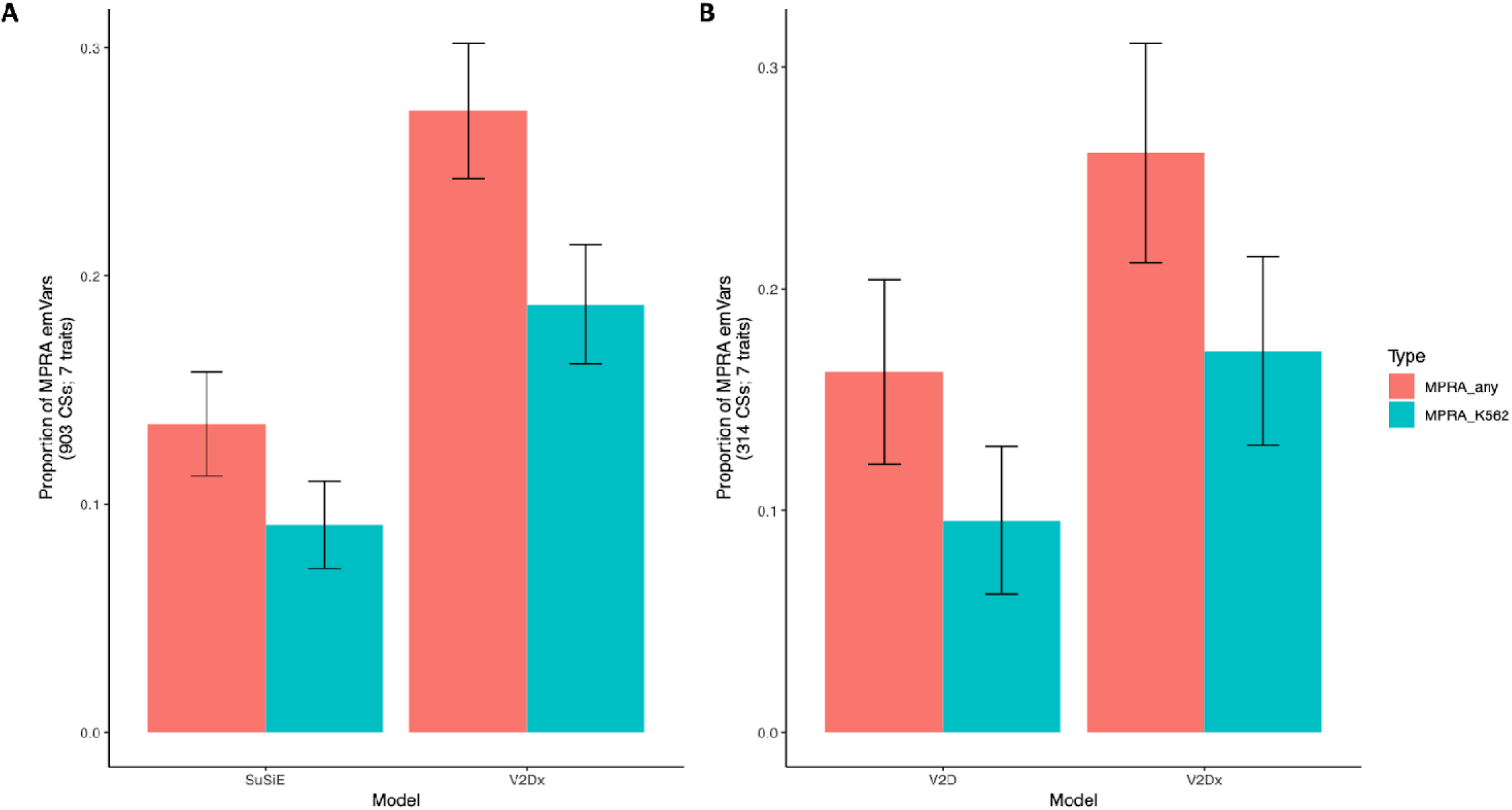
MPRA results for discordant credible sets in fine-mapping analyses of seven blood-related UK Biobank traits. We report the fraction of lead SNPs (variants with the highest PIP) that are expression-modifying variants (emVars) in credible sets with different lead SNPs from SuSiE and SuSiE+V2Dx (**A**) and SuSiE+V2D and SuSiE+V2Dx (**B**). Variants prioritized by SuSiE-V2Dx were significantly more likely to be emVars than variants prioritized by SuSiE (27.2 ± 1.5% vs. 13.5 ± 1.1% across 903 discordant CS; *P* = 6.1 x 10^-13^) and SuSiE+V2D (26.1 ± 2.4% vs. 16.2 ± 2.0% across 314 discordant CS; *P* = 0.005). Also, variants prioritized by SuSiE-V2Dx were significantly more likely to be K562 emVars than variants prioritized by SuSiE (18.8 ± 1.3% vs. 13.5 ± 1.1% across 903 discordant CS; *P* = 8.5 x 10^-9^) and SuSiE+V2D (17.2 ± 2.1% vs. 9.5 ± 1.6% across 314 discordant CS; *P* = 0.009).

## Supplementary Tables caption

**Table S1: List of the 15 UK Biobank independent GWAS.** These 15 traits correspond to the 16 independent traits from Weissbrod et al. 2020 Nat Genet, from which we removed hair color because of low polygenicity.

**Table S2: List of the 187 functional and evolutionary annotations from our baseline-LF models.** We report the 187 annotations of the baseline-LF model. They include 96 annotations for common variants (identical annotations from the baseline-LD model) and 91 annotations for low-frequency variants. We highlight 40 main common annotations used in **Fig. 2** and **Fig. 3**.

**Table S3: List of the 79 independent European GWAS not correlated with the 15 UK Biobank traits.** We report the set of 79 independent European GWAS constructed by removing from the 107 independent traits from ref. ^48^ the traits that were genetically correlated to the 15 UK Biobank GWAS.

**Table S4: Results of the simulations under the baseline-LD model.** We report true and estimated heritability functional enrichments for 40 representative functional annotations of the baseline-LD model.

**Table S5: Results of the simulations using an interactive effect between the coding and conserved annotations.** We report true and estimated heritability functional enrichments obtained with SuSiE and with S-LDSC without and with the interaction modeled (S-LDSC and S-LDSC+interaction, respectively).

**Table S6: Estimates of heritability enrichment estimated across 15 UK Biobank traits with S-LDSC, SuSiE-noprior, and SuSiE-prior.** We report heritability functional enrichments for 40 representative functional annotations of the baseline-LD model. S-LDSC values were averaged across the 15 traits.

**Table S7: Estimates of heritability enrichment across 15 UK Biobank traits estimated with S-LDSC, SuSiE-noprior, and SuSiE-prior.** We report heritability functional enrichments for 5 LD score bins. S-LDSC values were averaged across the 15 traits.

**Table S8: Mean squared error (MSE) of decision trees for common and low-frequency variants.** We report MSEs computed using a LEOCO procedure on common variants and low-frequency variants. For interpretability, we multiplied *b*^2^ by 10**7 and subtracted the MSE obtained by using the linear model (positive values mean that the decision tree does not provide a better fit than the linear model).

**Table S9: Heritability explained by each leaf of the common variant tree.** We report the common variant *h*^2^ enrichment of each leaf estimated by S-LDSC and expected by S-LDSC with the baseline-LD model using 79 independent European GWAS not correlated with the 15 UK Biobank traits.

**Table S10: Heritability explained by each leaf of the low-frequency variant tree.** We report the low-frequency variant *h*^2^ enrichment of each leaf estimated by S-LDSC and expected by S-LDSC with the baseline-LF model using 23 independent UK Biobank traits with sufficient power to investigate low-frequency variant architecture.

**Table S11: Benchmarking V2D scores across machine learning models.** We report *h^2^* enrichment computed on 79 independent GWAS and excess overlap computed across common variants fine-mapped with high confidence in MVP and FinnGen for V2D scores obtained using different machine learning methods.

**Table S12: MSE of machine learning models for common and low-frequency variants.** We report MSE computed using a LEOCO procedure on common variants and low-frequency variants. For interpretability, we multiplied *b*^2^ by 10**7 and subtract the MSE obtained by using the linear model (positive values mean that the model does not provide a better fit than the linear model).

**Table S13: Benchmarking V2D scores across existing prioritization scores.** We report heritability enrichment computed on 79 independent GWAS and excess overlap computed across common variants fine-mapped with high confidence in MVP and FinnGen for V2D scores obtained using existing prioritization scores.

**Table S14: SuSiE+V2D results across 110 UK Biobank traits.** We report SNPs with PIP > 0.50.

**Table S15: SuSiE results across 110 UK Biobank traits.** We report the number of SNPs with PIP greater than 0.5, 0.75 and 0.9.

**Table S16. Expression-modifying variants (emVars) identified by SuSiE versus SuSiE+V2D.**

**Table S17. Expression-modifying variants (emVars) identified by SuSiE+cV2F-bLD versus SuSiE+V2D.**

